# Cannabis use and atherosclerotic cardiovascular disease: a Mendelian randomization study

**DOI:** 10.1101/2023.02.23.23286339

**Authors:** Roxane de La Harpe, Tabea Schoeler, Christian W Thorball, Aurélien Thomas, Zoltán Kutalik, Julien Vaucher

## Abstract

**Background:** Research on the link between cannabis use and the development of atherosclerotic cardiovascular disease (ASCVD) is inconsistent and challenging to interpret, given existing study limitations.

**Aims:** To estimate the effects of genetically indexed cannabis use on the risk of coronary artery disease (CAD) and acute ischemic stroke (IS).

**Methods:** 65 independent single-nucleotide polymorphisms (SNPs), obtained from a genome-wide association study on lifetime cannabis use (n=184,765), were employed as instruments to estimate the association between genetically indexed cannabis use and risk of CAD and IS using a two-sample Mendelian Randomization (MR) approach. Summary statistics on CAD (CARDIoGRAMplusC4D Consortium; 60,801 cases and 123,504 controls) and IS (MEGASTROKE; 34,217 cases and 406,111 controls) were obtained separately. A comprehensive review of the observational literature on cannabis use and CAD or IS was also performed and contrasted with MR results.

**Results:** There was no causal effect of cannabis use on the risk of CAD (odds ratio (OR) per ever-users vs. never-users 0.93; 95% confidence interval (CI), 0.83 to 1.03) or IS (OR 1.05; 95%CI, 0.93 to 1.19). Sensitivity analyses yielded similar results, and no heterogeneity and directional pleiotropy were observed. Our meta-analysis of observational studies showed no significant association between ever use of cannabis with risk of CAD (k=6 studies; OR_pooled_=1.23, 95% CI 0.78 to 1.69), nor with IS (k=6 studies; OR_pooled_=1.22, 95% CI 0.95 to 1.50).

**Conclusion:** Using a genetic approach approximating a clinical trial revealed no evidence for a causal effect of genetic predisposition to cannabis use on CAD or IS development. These findings are reassuring from a public health perspective, as ever cannabis use is unlikely to contribute to the risk of ASCVD.

## Introduction

Cannabis is one of the most psychotropic substances used globally, with almost 4% of the population aged 15–64 years having consumed cannabis at least once in 2021.[1] More evidence on the impact of cannabis use on health is thus necessary at population-wide and individual levels, especially with atherosclerosis cardiovascular diseases (ASCVD) accounting for 30% of globally deaths. While an association between cannabis use and risk of atherosclerosis cardiovascular diseases (ASCVD) has been reported numerous times [2–4], it remains inconclusive as to whether this link is causal in nature. In experimental studies, Cannabidiol (CBD) and tetrahydrocannabinol (THC), substances both present in cannabis, have been found to have potential beneficial effects against ASCVD development, mainly through anti-oxidative and anti-apoptotic effects.[5–13] Conversely, evidence also points towards adverse cardiovascular effects of THC, such as a decrease in myocardial contractility, vasospasm, tachycardia and systolic blood pressure increase, conditions that are known to promote ASCVD development.[14–16]

Causality between cannabis use and ASCVD is challenging to assess in observational studies due to recall bias, inadequate exposure assessment, non-exhaustive inclusion of confounders or weak methodology design.[17] Since a deliberate and long-term exposure to cannabis would be unethical, a clinical trial, removing potential biases (e.g., confounding or reverse causation) in the cannabis-ASCVD association, is not possible. A genetic approach, recapitulating a randomized trial, thus represents an asset to infer a causal association between a potential harmful exposure (cannabis) and a disease outcome (ASCVD).[18] Recently, Zhao et al, using Mendelian randomization (MR) principles, did not find a causal association between cannabis use and ASCVD, but showed some evidence for a causal effect of cannabis use on small vessel stroke and atrial fibrillation.[19] However, their MR analysis was based solely on 10 genetic instruments, since it was not derived from the largest and most recent genome-wide association study (GWAS) of cannabis use, which may have reduced the statistical power of their analysis.

To obtain more reliable results, here we used 65 independent genetic markers (from the most recent GWAS of cannabis use) to perform Mendelian randomization analyses. We included several sensitivity analyses and tested for the presence of pleiotropic effects of the instruments to ensure robust causal association results between cannabis consumption and both coronary artery disease (CAD) and ischemic stroke (IS). Further, we assessed whether adjusting for genetically indexed tobacco use altered the association. Finally, we performed a meta-analysis of published observational studies and contrasted the result with the causal estimates.

## Methods

### Principles of two-sample Mendelian Randomization

Mendelian randomization (MR) is a statistical method using measured variation in single-nucleotide polymorphisms (SNPs) associated with an exposure to examine the causal effect of this exposure (cannabis use) on a disease outcome (CAD or IS). SNPs, used as genetic instruments, have to meet three assumptions to be valid instruments: i. relevance assumption (genetic instruments have to be robustly associated with the exposure of interest); ii. independence assumption (they should not be associated with any confounder of the exposure-outcome relationship); iii. exclusion restriction assumption (they can affect the outcome only through the exposure).[20,21]

Two-sample MR refers to the application of MR to summary genetic statistics estimated in two non-overlapping sets of individuals. The “first” sample is used for computing a genetic instrument for the exposure. The “second” sample is employed to estimate the instrument-outcome association. These two associations are then used to estimate the underlying causal effect[20].

### Genetic markers associated with ever use of cannabis

We used a publicly available GWAS computed from three distinct sources (ICC study, UK-Biobank and 23andMe), with a combined sample size of 184,765 participants of European ancestry, on ever use of cannabis (including 53,179 cases, 131,586 controls) (Supplementary Table 1 for details about studies included).[22] All alleles in the GWAS were reported from the positive strand. Pasman et al. executed linkage disequilibrium clumping to eliminate genetically correlated SNPs (R^2^<0.001) and proposed 69 independent SNPs linked to ever use of cannabis, explaining 1.12% of the variance in cannabis use. Among these, we excluded four SNPs (rs11749751, rs2335349, rs3740390 and rs61942416) with discordant direction of effect among the three sources. We confirmed the independence of the SNPs with the SNP Annotation and Proxy Search tool (SNAP, Broad Institute, MA, US https://data.broadinstitute.org/mpg/snpsnap/; as accessed on 2022, 4 Dec) (Supplementary Table 4). We selected then 64 SNPs; 5 which surpassed the conventional genome-wide significance threshold for genome-wide association with lifetime cannabis use (p-value <5×10^−8^) and 59 other SNPs that passed a more lenient significance threshold (p-value <5×10^−5^), but could be considered as an additional instrumental variable for the MR analysis (Supplementary Table 2).[23]

### Genetic markers associated with ASCVD

No publicly available GWAS repository on ASCVD was found. We, therefore, assessed the instrument-outcome association separately for CAD, using the Coronary Artery Disease Genome-wide Replication and Meta-analysis plus the Coronary Artery Disease 1000 Genomes-based GWAS (CARDIoGRAMplusC4[24]) and IS, using the Multiancestry GWAS with stroke and stroke sub-types (MEGASTROKE[25]).

CARDIoGRAMplusC4D Consortium (www.cardiogramplusc4d.org/) involved 60,801 cases and 123,504 controls from 39 studies in a GWAS meta-analysis of CAD with 77% of European ancestry and about 70% of acute myocardial infarction (AMI). MEGASTROKE (www.megastroke.org) included 34,217 European cases and 406,111 European controls from 16 studies in a GWAS of IS.

The 64 SNPs associated with ever use of cannabis were matched and harmonized (i.e. matching the reference alleles) across the data sets. One SNP (rs80144387) was not available in CARDIoGRAMplusC4 data and was thus excluded from the analyses (Supplementary Table 2).

There was no overlap between participants from the GWAS of cannabis use and MEGASTROKE. Only two studies, including 3,735 participants, contributed in both GWAS of cannabis use and CARDIoGRAMplusC4D (Supplementary Table 1 and supplementary Figure 1).

### Observational association between ever use of cannabis and ASCVD

There was no meta-analysis in the literature reporting pooled observational association (correlation) estimates between cannabis use and ASCVD or, separately, for CAD or IS. We, therefore, conducted a random-effect meta-analysis, including studies assessing the association between cannabis use and ASCVD. Among the studies identified in a comprehensive literature search, we selected only prospective and retrospective observational studies. In addition, we only used studies that reported ever use of cannabis (compared with never users) as an exposure and a corresponding risk estimate (expressed as odds ratio (OR) or hazard ratio (HR)) for ASCVD, CAD or IS. Supplementary Figure 2 presents the literature search strategy and supplementary Tables 3 and 4 summarize the main characteristics of included and excluded studies, respectively.

### Statistical analysis

MR was conducted, using Stata v.17 (Stata, College Station, TX, USA, using mrobust package, available at: https://github.com/remlapmot/mrrobust) and R Statistical Software (v4.1.2; R Core Team 2021, using TwoSampleMR package v0.5.6, available at: https://mrcieu.github.io/TwoSampleMR/articles/index.html). Analyses were performed for CAD and IS separately.

We first generated a causal estimate for each instrumental variable (i.e., SNPs) by dividing the association of each SNP with risk of CAD/IS by the corresponding association with risk of ever use cannabis. The standard error (s.e.) was estimated using the delta method.[26] We then pooled together the individual causal effect estimates using fixed-effects (inverse variance weighted [IVW]) meta-analysis. As a sensitivity analysis, we also pooled together estimates using random-effects meta-analysis. To compare the pooled causal estimates to the pooled observational estimates, we transformed the summary estimates from meta-analysis into “users vs. non-users” of cannabis, as opposed to a per-1-log-unit increase in ever use of cannabis. We used estimates for the risk of CAD/IS in the general population, and the prevalence of CAD/IS among never users of cannabis, as previously described[27]. A full description of the methodology is described in the Supplementary Methods. We also conducted a Steiger filtering analysis to test the direction of the causal estimate. This approach assumes that a valid instrumental variable should explain more variance for the exposure than for the outcome and identifies SNPs that do not satisfy this criterion (SNP-outcome correlation greater than the SNP-exposure correlation).[28]

### Strength of genetic instruments and power to detect a causal effect

We estimated instrument strength by calculating the proportion of variance in ever use of cannabis explained by each SNP. We used then F-statistic for each SNP individually and cumulatively assuming that F-statistics >20 represents an acceptable correlation.[29] In the present study, the cumulative F-statistic was 29.1 minimizing the risk of weak instrument bias. Full details are provided in the Supplementary Methodology and supplementary Table 2.

The power of our MR analysis to detect the same magnitude of association reported in the observational studies, using a two-sided α of 0.05, was 98% for both CAD and IS (Supplementary Table 5).

### Assessment of horizontal pleiotropy

To test the robustness of the causal estimation, we tested for the presence of pleiotropy. Egger Mendelian randomization (MR-Egger) method detects and corrects for the bias due to directional pleiotropy, allowing one or more SNPs to have pleiotropic effects, as long as the size of these pleiotropic effects is independent of the size of the SNPs effects on the exposure.[30] The methodology resembles conventional MR analysis (IVW), except that the intercept of the weighted linear regression is unconstrained (opposite as constrained equal to zero in IVW method).[30] A low p-value for the MR-Egger intercept test suggests pleiotropy. The s.e. was obtained by bootstrap resampling 10’000 times. Finally, the *I*^*2*^ statistic in the context of MR-Egger quantifies weak instrument bias and was low in our analysis (*I*^*2*^=17% for CAD; and *I*^*2*^=39% for IS).

We then applied simulation extrapolation (MR-Egger-SIMEX implemented in Stata using the mrrobust package) to adjust the MR-Egger causal estimates to account for a potential NOME violation (NO Measurement Error assumption, the assumption that the SNP-exposure association is true).[31]

We also conducted a weighted median MR analysis (implemented in Stata using the mrobust package), which gives more weight to SNPs with homogeneous causal estimates (that is, close to the median causal estimate) even when up to 50% of the weight in the analysis arises from invalid SNPs.[32]

### Sensitivity analysis

Tobacco consumption is a risk factor for ASCVD and shares a strong genetic correlation with use of cannabis.^25,26^ We then conducted a multivariable analysis adjusting for SNP-tobacco, to adjust for shared pathways with and/or potential confounding by tobacco, using summary statistics for the association of each of the 64 cannabis-related SNPs with tobacco. Smoking status was derived from 1,232,091 European individuals with 557,337 ever smoker phenotypes (vs never smoker) in the GWAS & Sequencing Consortium of Alcohol and Nicotine (GSCAN Consortium, https://conservancy.umn.edu/handle/11299/201564[35] downloads). Multivariable MR was conducted by regressing the SNP–cannabis estimates on SNP–CAD or -IS estimates adjusting for SNP–tobacco estimates. The s.e. was obtained by bootstrap resampling 10 000 times. Eight of 64 cannabis-related SNPs were not available in the GSCAN Consortium and therefore excluded from this analysis.

We finally computed three sensitivity analyses to test the coherence of our results. First, we restricted the level of genome-wide significance by the selection of genetic variants with a p-value<5×10^−8^ (Supplementary Table 2). Second, as it was not possible to verify that the alleles reported by CARDIoGRAMplusC4D or MEGASTROKE have been correctly orientated, we selected SNPs in low linkage-disequilibrium with other SNPs (r2 < 0.001) within a clumping distance of 10,000 kb and removed palindromic SNPs if the allele frequency was close to 50%.[36] Third, as “ever use of cannabis” phenotype and genetic instruments derived from it can suffer from a lack of specificity, we repeated the analysis using SNPs from a recent GWAS of cannabis use disorder, comprising 14,080 cases and 343,726 controls of unrelated individuals from European ancestry (Psychiatric Genomic Consortium, https://pgc.unc.edu/for-researchers/download-results/downloads, more details in Supplementary Methods).[37]

## Results

### Observational association between ever use of cannabis and risk of CAD and IS

Twelve studies met our primary research criteria. Six reported ever use of cannabis (compared with no use) along with CAD status and six others measured ever use of cannabis along with IS. When meta-analysing these estimates, ever use of cannabis was not significantly associated with risk of CAD using random-effects modelling (OR_pooled_ 1.23, 95% confidence interval (CI) 0.78 to 1.69, I^2^ 90.8%; Supplementary Figure 3), nor with risk of IS (OR_pooled_ 1.22, 95% CI 0.95 to 1.50, I^2^ 85.4%; Supplementary Figure 4). Combining all 12 studies together (encompassing 714,938 cases and 144 million controls), ever use of cannabis was significantly associated with overall ASCVD (OR_pooled_ 1.27, 95% CI 1.03 to 1.51, p-value<0.001, I^2^ 97.3%), using a random-effects modelling (Figure 1). The cannabis-ASCVD association was similar when only prospective studies were selected (Supplementary Figure 5). Egger’s test via funnel plot asymmetry, as measured a linear regression of the effect estimates on their standard errors weighted by their inverse variance, was not significant (0.64, 95% CI −0.54 to 1.81, p-value=0.3), indicating an unlikely publication bias.

**Figure 1.**
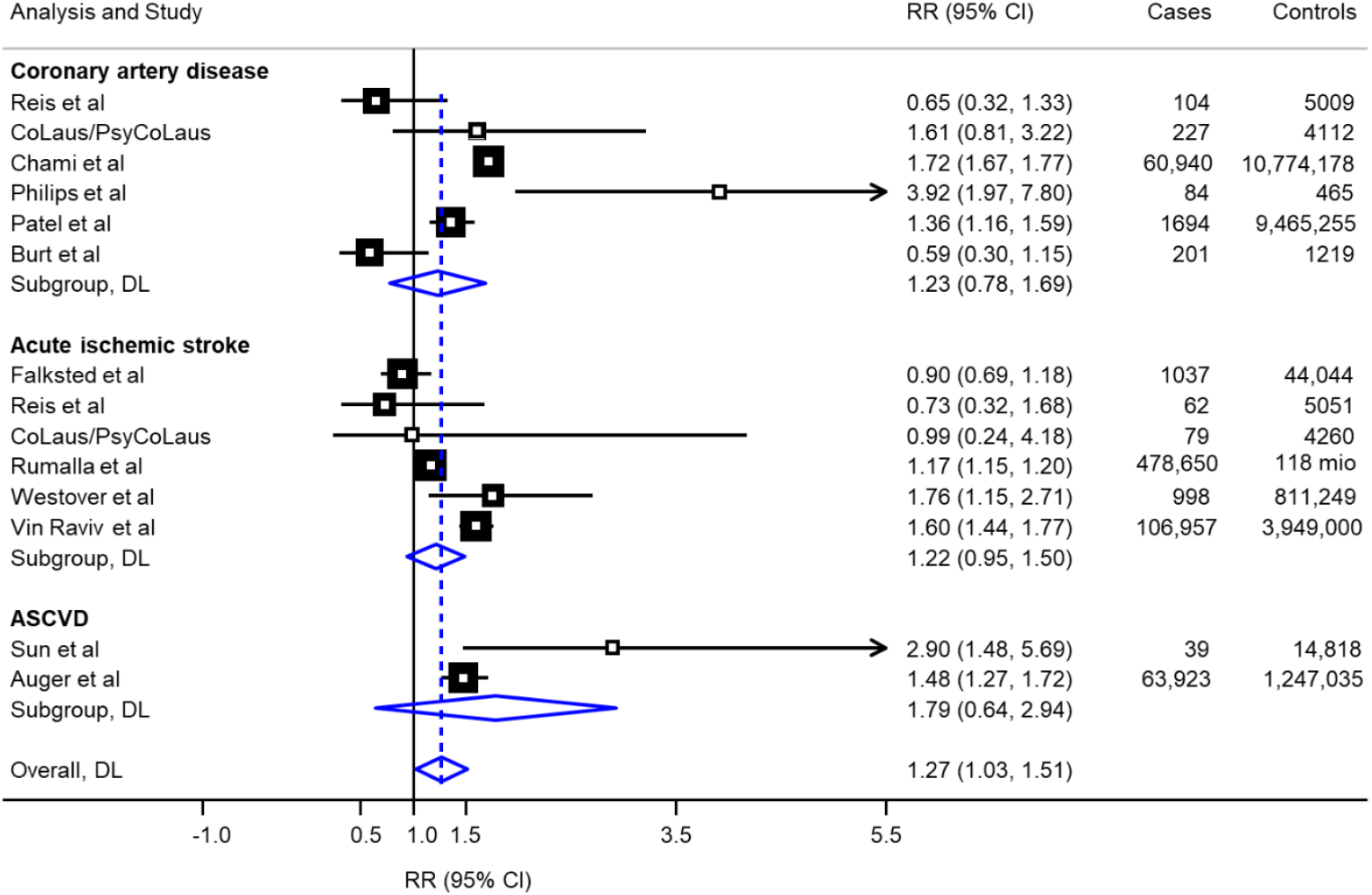
Meta-analysis of observational studies reporting an association between use of cannabis and risk of atherosclerotic cardiovascular disease. Meta-analysis uses a random-effects model, DerSimonian and Lair methods (DL). Studies are sorted by type of outcome (coronary artery disease, ischemic stroke or global ASCVD analysis). Relatives risk (RR) and 95% confidence intervals (CI) express the risk of ASCVD for “ever use of cannabis” (compared with never use). For additional information on each study, see Supplementary Table 1. Supplementary Figure 4 and 5 provide meta-analysis stratified by outcome and type of risk ratio.

### Causal effect estimates of ever use of cannabis on risk of CAD and IS

The 64 SNPs associated with ever use of cannabis explained 1% of its variance. In MR analysis, ever use of cannabis was not causally associated with risk of CAD (OR per-1-log unit in ever use of cannabis [derived by fixed-effect meta-analysis of individual causal effects estimates of SNPs], 0.97, 95% CI 0.92 to 1.02, p-value=0.19; Figure 2.A and Supplementary Figure 6). Similarly, no causal effect was found when IS was assessed as the outcome (OR per-1-log unit in ever use of cannabis, 1.03, 95% CI 0.98 to 1.09, p-value=0.41; Figure 2.B and Supplementary Figure 7). Random-effects meta-analysis showed converging results (OR 0.97, 95% CI 0.92 to 1.02, p-value=0.19 for CAD and 1.03, 95% CI 0.97 to 1.09, p-value=0.38 for IS). Steiger filter showed correct causal direction for both CAD and IS overall (p-value<0.001 for both) and for each SNP, separately.

**Figure 2.**
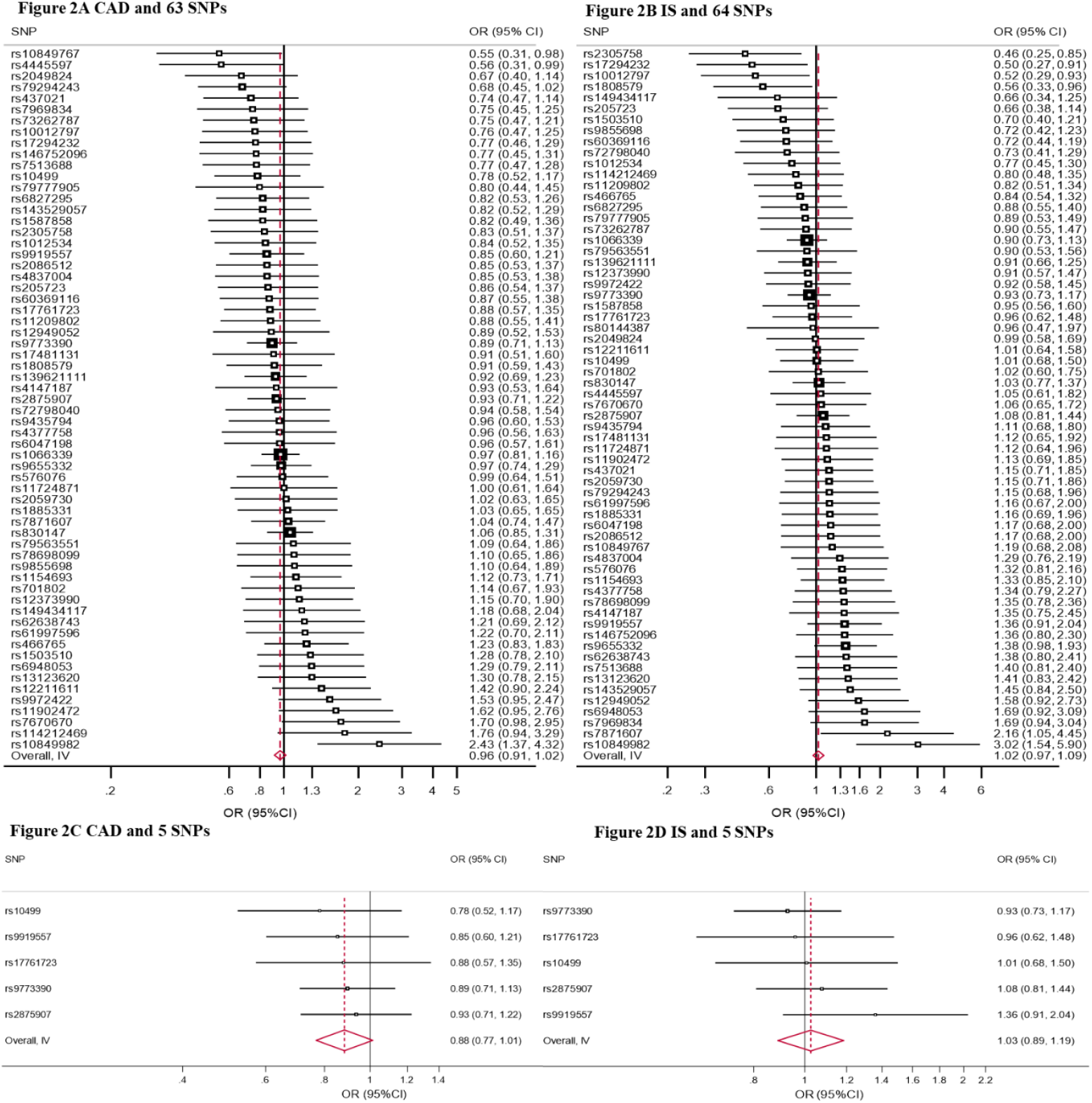
Forest plot showing the effect size of lifetime cannabis use on risk of CAD (**Figure 2A**) and of IS (**Figure 2B**) for each cannabis use-associated genetic marker and when using a more stringent threshold for selection of the SNPs (p<10^e-8^) for CAD (**Figure 2C**) and for IS (**Figure 2D**) Meta-analysis of the association of genetically instrumented use of cannabis and risk of CAD for the 63 single-nucleotide polymorphisms (SNPs) (Figure 2A) and of IS for the 64 SNPs (Figure 2B). The second meta-analysis shows results when using 5 SNPs which surpassed the conventional genome-wide significance threshold for genome-wide association with lifetime cannabis use (p-value <5×10^−8^) for CAD (Figure 2C) and for IS (Figure 2D). Odds ratios (OR) and 95% confidence intervals (CI) express the risk of event per-1-log unit increase in ever use of cannabis. Meta-analysis uses a fixed effect model.

The MR estimate transformed in population-based OR (OR per users vs. non-users for CAD, 0.93, 95% CI 0.83 to 1.03; Figure 3 and OR for IS, 1.05, 95% CI, 0.93 to 1.19; Figure 4) was consistent with estimates derived from observational analysis for CAD (test for heterogeneity between group, p-value=0.185) and IS (test for heterogeneity between group, p-value=0.053).

**Figure 3.**
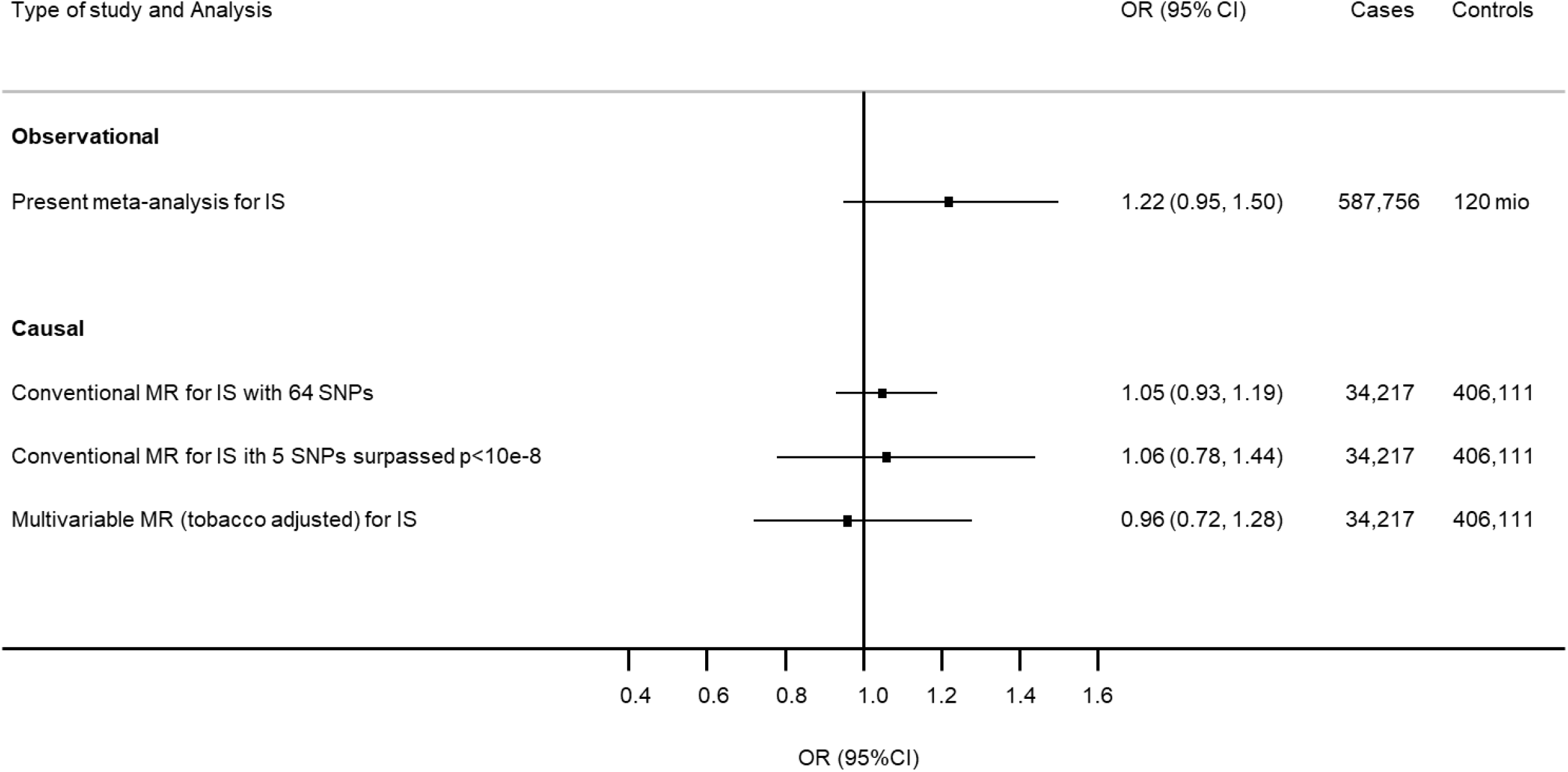
Comparison of observational and causal estimates for use of cannabis and risk of CAD Observational estimates are provided according to the meta-analysis reported in Figure 1 restricted to coronary artery disease, as separate outcome for ever use of cannabis. Causal estimates represent population-based association derived by conventional (Figure 2) and multivariable Mendelian randomization. The method to derive the population-based OR of ASCVD among users of cannabis compared with non-users is described in the Supplementary Methods.

**Figure 4.**
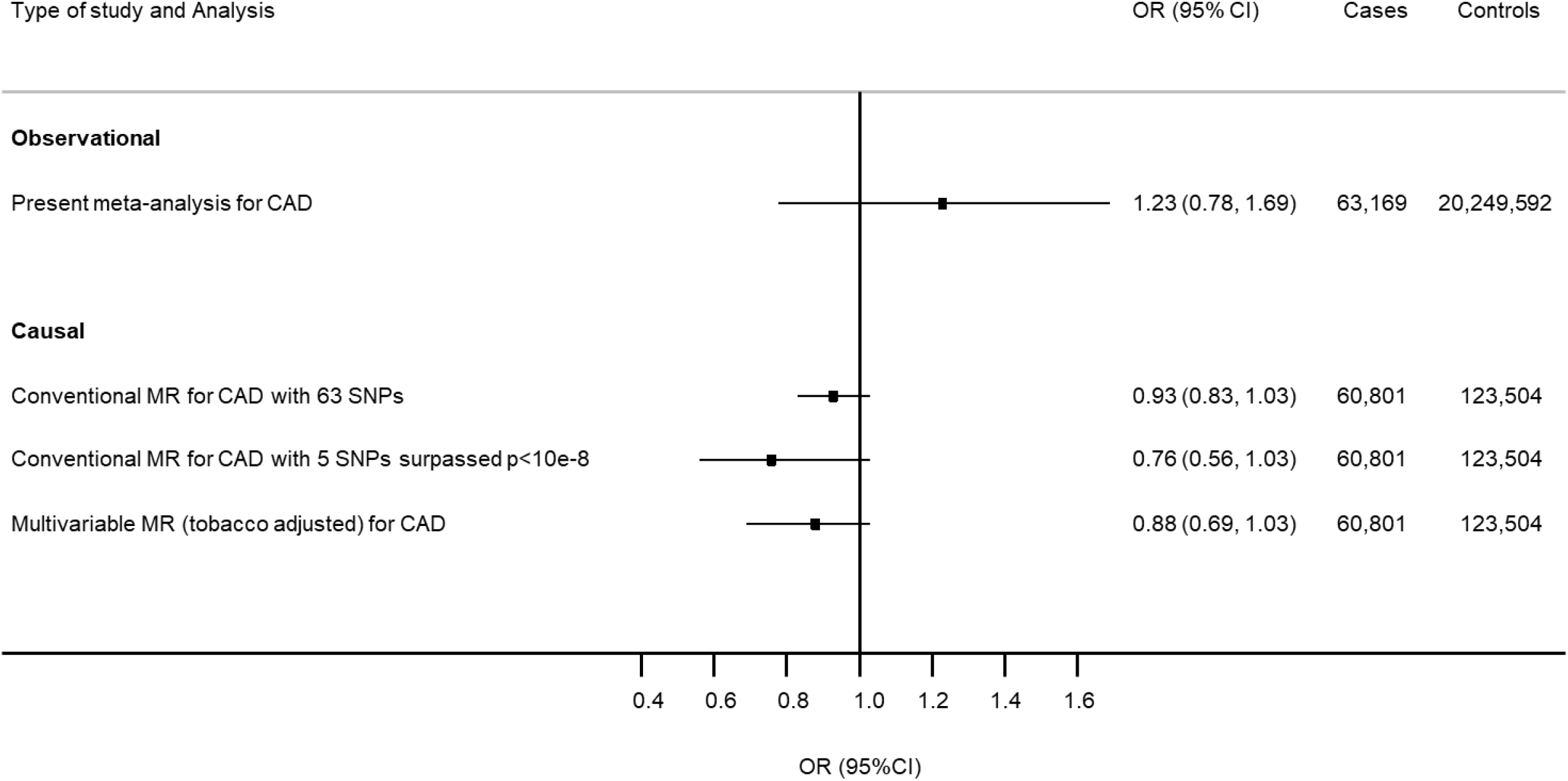
Comparison of observational and causal estimates for use of cannabis and risk of IS Observational estimates are provided according to the meta-analysis reported in Figure 1 restricted to acute ischemic disease, as separate outcome for ever use of cannabis. Causal estimates represent population-based association derived by conventional (Figure 2) and multivariable Mendelian randomization. The method to derive the population-based OR of ASCVD among users of cannabis compared with non-users is described in the Supplementary Methods.

### Assessment of pleiotropic effects of the genetic markers

We did not find evidence against the null hypothesis of no directional pleiotropy of the genetic markers using MR-Egger (P-value for pleiotropy of 0.766 for CAD and 0.653 for IS). The causal estimates derived from MR-Egger, MR-Egger adjusted for simulation extrapolation (SIMEX) and weighted median MR produced consistent causal estimate compared with conventional MR estimates for CAD (Supplementary Table 6 and Supplementary Figure 8) and IS (Supplementary Table 7 and Figure 9).

### Sensitivity analysis

Adjusting for smoking in multivariable MR did not show evidence of shared pathways and/or confounding with a causal effect estimate of CAD and IS (OR per-1-log unit 0.94, 95% CI 0.84 to 1.05 for CAD; and 0.97, 95% CI 0.86 to 1.12 for IS, ORs per users vs. non-users are shown in Figure 3 and 4). The pattern of the MR estimates did not change after using a more stringent threshold for the selection of genetic variants (p-value <5×10^−8^) (Figure 2, 3 and 4 and Supplementary Table 8). The effect was consistent with a more conservative approach selecting genetic instruments (removal of palindromic SNP with intermediate minor allele frequency) (Supplementary Table 9), or when we estimated the causal effect for cannabis use disorder as exposure (Supplementary Table 10).

## Discussion

Using a genetically informed causal inference approach, this study provides no evidence that genetically indexed cannabis use has a causal effect on CAD or IS risk, a result robust to a range of sensitivity analyses. This finding is in line with previous MR results relying on less powerful genetic instruments.[19] They are also consistent with a meta-analysis which did not find serious cardiovascular events in a randomized controlled trial (RCT) using medical cannabinoids.[38] These findings are important from individual- and public health perspectives, considering the increase in prevalence of medical and recreational use of cannabis.[1]

Our meta-analysis, combining estimates obtained in observational studies, showed no significant association between cannabis use and CAD or IS development individually, which is coherent with our MR findings. We found a significant association between cannabis use and overall ASCVD development. Whereas this result was not affected by publication bias, there was substantial levels of heterogeneity between studies, which can lead to this spurious significant association between cannabis use and overall ASCVD development.

Our study shows no association between cannabis use and atherosclerotic cardiovascular disease, despite numerous observational studies reporting a detrimental association.[2–4,39–42] Differences in cannabis doses, formulations, exposition-time and pattern of use can have different and divergent effects and contributions on the occurrence of different ASCVDs and thus lead to confusion when testing their contribution together and for an overall ASCVD outcome. Indeed, cannabis use exposed to hundreds of cannabinoids, the effects of most of which are not known, especially with regard to their affinity to cannabinoid receptor (CBR). Even the two main cannabinoids, namely cannabidiol (CBD) and tetrahydrocannabinoid (THC), involved in the use of cannabis have different known or hypothesized effects of the two substances involved in the use of cannabis. The endocannabinoid system includes two receptors cannabinoid receptor 1 (CBR1) and 2 (CBR2) with different biological roles.[12] THC is an agonist of the CBR1 receptor, which, via the autonomic nervous system, induces an increase in heart rate and blood pressure [12,43], thus suggested as a trigger for ASCVD in case of predisposition.[42] Conversely, high-doses of THC, translate into a decrease<u>s</u> in heart rate, as well as blood pressure with a decrease in cerebral flow, which is suggested as a mechanism for the development of IS.[43] Activation of CBR2 has been shown to regulate inflammation and may limit the production of oxidized lipoprotein by modulating the effect of CBR1 in the development of atherosclerosis. Whereas the mechanisms of action of CBD, particularly in relation to inflammation, remain obscure, indirect effects on anandamide which will modulate CBR1 and CBR2 have been considered for [5,7,11,12,43]. Therefore, further MR studies with SNPs specific to THC, CBD use or CBR1/CBR2 agonist or antagonist could distinguish the effects of the two main substances composing cannabis.

Weak instrumental variables (that did not achieve GWAS significance) can lead to downward bias and hence loss in statistical power.[44] In addition, Winner’s curse can yield downward biased causal effect estimates when the exposure and outcome data sets have little overlap, like ours.[45],[46] The availability of only a few SNPs reaching the conventional genetic significance threshold of <5×10^−8,^, further reduces power. However, the F-statistic provided evidence against weak instrumental bias. Other limitations include that our study did not allow the investigation of the risk of ASCVD in relation to the quantity, type, route of administration, or the age at exposure to cannabis. Second, all genetic summary statistics are from European ancestry, except for the CardioGRAMplusC4D (23% of participants from a different ethnical background). Differences in ancestry may mean, for example, that a genetic association with cannabis may be true in that specific population, but, due to differences in linkage disequilibrium, this will not be the case in a different ancestry group, which in turn can affect the causal association with ASCVD.

### Conclusion

Our genetic approach, approximating a randomized control trial that would be unethical in these circumstances, showed no evidence for a causal effect of genetic liability to cannabis use on risk of CAD or IS. Knowing the burden of ASCVD and the frequency of cannabis use in the general population, these findings are reassuring, as cannabis use does not likely participate to the development of ASCVD.

## Data Availability

This study included summary statistics of different publicly available sources. First, a genetic study on cannabis use (from Pasman et al, in press Nature Neuroscience), which would like to acknowledge all participating groups of the International Cannabis Consortium, and in particular the members of the working group including Joelle Pasman, Karin Verweij, Nathan Gillespie, Eske Derks, and Jacqueline Vink. Pasman et al. Then, data on coronary artery disease and myocardial infarction have been contributed by CARDIoGRAMplusC4D investigators and have been downloaded from www.CARDIOGRAMPLUSC4D.ORG.
Data on stroke came from The MEGASTROKE project who received funding from sources specified at http://www.megastroke.org/acknowledgments.html.
Then, a genetic study on smoking (from Liu et al, in press Nature Genetics) available at https://conservancy.umn.edu/handle/11299/2015.
Finaly, some data from The Substance Use Disorders Working Group of the Psychiatric Genomics Consortium (PGC-SUD) which is supported by funds from NIDA and NIMH to MH109532 and gratefully acknowledge the participants in those studies without whom this effort would not be possible.

https://www.ru.nl/bsi/research/group-pages/substance-use-addiction-food-saf/vm-saf/genetics/international-cannabis-consortium-icc/

http://www.cardiogramplusc4d.org/data-downloads/

https://www.megastroke.org/

https://conservancy.umn.edu/handle/11299/2015

https://www.med.unc.edu/pgc/download-results/

## Reference

1. WDR 2021_Booklet 3 [Internet]. United Nations: Office on Drugs and Crime. [cited 2022 Feb 3]. Available from: http://www.unodc.org/unodc/en/data-and-analysis/wdr-2021_booklet-3.html

2. Jouanjus E, Raymond V, Lapeyre-Mestre M, Wolff V. What is the Current Knowledge About the Cardiovascular Risk for Users of Cannabis-Based Products? A Systematic Review. Curr Atheroscler Rep. 2017 Jun;19(6):26.

3. Skipina TMS, Patel NP, Upadhya BU, Soliman EZS. Cannabis use is associated with prevalent coronary artery disease. European Heart Journal. 2021 Oct 1;42(Supplement_1):ehab724.1258.

4. Sun Y, Liu B, Wallace RB, Bao W. Association of Cannabis Use With All-Cause and Cause-Specific Mortality Among Younger- and Middle-Aged U.S. Adults. Am J Prev Med. 2020 Dec;59(6):873–9.

5. Steffens S, Veillard NR, Arnaud C, Pelli G, Burger F, Staub C, Karsak M, Zimmer A, Frossard JL, Mach F. Low dose oral cannabinoid therapy reduces progression of atherosclerosis in mice. Nature. 2005 Apr 7;434(7034):782–6.

6. Shmist YA, Goncharov I, Eichler M, Shneyvays V, Isaac A, Vogel Z, Shainberg A. Delta-9-tetrahydrocannabinol protects cardiac cells from hypoxia via CB2 receptor activation and nitric oxide production. Mol Cell Biochem. 2006 Feb;283(1–2):75–83.

7. Durst R, Danenberg H, Gallily R, Mechoulam R, Meir K, Grad E, Beeri R, Pugatsch T, Tarsish E, Lotan C. Cannabidiol, a nonpsychoactive Cannabis constituent, protects against myocardial ischemic reperfusion injury. Am J Physiol Heart Circ Physiol. 2007 Dec;293(6):H3602–3607.

8. England TJ, Hind WH, Rasid NA, O’Sullivan SE. Cannabinoids in experimental stroke: a systematic review and meta-analysis. J Cereb Blood Flow Metab. 2015 Mar;35(3):348–58.

9. Shayesteh MRH, Haghi-Aminjan H, Mousavi MJ, Momtaz S, Abdollahi M. The Protective Mechanism of Cannabidiol in Cardiac Injury: A Systematic Review of Non-Clinical Studies. Curr Pharm Des. 2019;25(22):2499–507.

10. Garza-Cervantes JA, Ramos-González M, Lozano O, Jerjes-Sánchez C, García-Rivas G. Therapeutic Applications of Cannabinoids in Cardiomyopathy and Heart Failure. Oxid Med Cell Longev. 2020;2020:4587024.

11. Huang W, Zeng Z, Lang Y, Xiang X, Qi G, Lu G, Yang X. Cannabis Seed Oil Alleviates Experimental Atherosclerosis by Ameliorating Vascular Inflammation in Apolipoprotein-E-Deficient Mice. J Agric Food Chem. 2021 Aug 18;69(32):9102–10.

12. Kicman A, Toczek M. The Effects of Cannabidiol, a Non-Intoxicating Compound of Cannabis, on the Cardiovascular System in Health and Disease. Int J Mol Sci. 2020 Sep 14;21(18):6740.

13. Mathew B, Harilal S, Musa A, Kumar R, Parambi DGT, Jose J, Uddin MS, Shah MA, Behl T, Unnikrishnan MK. An Agathokakological Tale of Δ9-THC: Exploration of Possible Biological Targets. Curr Drug Targets. 2021;22(7):823–34.

14. Bonz A, Laser M, Küllmer S, Kniesch S, Babin-Ebell J, Popp V, Ertl G, Wagner JA. Cannabinoids acting on CB1 receptors decrease contractile performance in human atrial muscle. J Cardiovasc Pharmacol. 2003 Apr;41(4):657–64.

15. Zubrzycki M, Liebold A, Janecka A, Zubrzycka M. A new face of endocannabinoids in pharmacotherapy. Part I: protective role of endocannabinoids in hypertension and myocardial infarction. J Physiol Pharmacol. 2014 Apr;65(2):171–81.

16. Franz CA, Frishman WH. Marijuana Use and Cardiovascular Disease. Cardiol Rev. 2016 Aug;24(4):158–62.

17. Gage SH, Jones HJ, Burgess S, Bowden J, Davey Smith G, Zammit S, Munafò MR. Assessing causality in associations between cannabis use and schizophrenia risk: a two-sample Mendelian randomization study. Psychol Med. 2017 Apr;47(5):971–80.

18. Davies NM, Holmes MV, Smith GD. Reading Mendelian randomisation studies: a guide, glossary, and checklist for clinicians. BMJ. 2018 Jul 12;362:k601.

19. Zhao J, Chen H, Zhuo C, Xia S. Cannabis Use and the Risk of Cardiovascular Diseases: A Mendelian Randomization Study. Front Cardiovasc Med. 2021;8:676850.

20. Pierce BL, Burgess S. Efficient design for Mendelian randomization studies: subsample and 2-sample instrumental variable estimators. Am J Epidemiol. 2013 Oct 1;178(7):1177–84.

21. Reading Mendelian randomisation studies: a guide, glossary, and checklist for clinicians | The BMJ [Internet]. [cited 2022 Feb 3]. Available from: https://www.bmj.com/content/362/bmj.k601

22. Pasman JA, Verweij KJH, Gerring Z, Stringer S, Sanchez-Roige S, Treur JL, Abdellaoui A, Nivard MG, Baselmans BML, Ong JS, Ip HF, van der Zee MD, Bartels M, Day FR, Fontanillas P, Elson SL, 23andMe Research Team, de Wit H, Davis LK, MacKillop J, Substance Use Disorders Working Group of the Psychiatric Genomics Consortium, International Cannabis Consortium, Derringer JL, Branje SJT, Hartman CA, Heath AC, van Lier PAC, Madden PAF, Mägi R, Meeus W, Montgomery GW, Oldehinkel AJ, Pausova Z, Ramos-Quiroga JA, Paus T, Ribases M, Kaprio J, Boks MPM, Bell JT, Spector TD, Gelernter J, Boomsma DI, Martin NG, MacGregor S, Perry JRB, Palmer AA, Posthuma D, Munafò MR, Gillespie NA, Derks EM, Vink JM. GWAS of lifetime cannabis use reveals new risk loci, genetic overlap with psychiatric traits, and a causal influence of schizophrenia. Nat Neurosci. 2018 Sep;21(9):1161–70.

23. Burgess S, Small DS, Thompson SG. A review of instrumental variable estimators for Mendelian randomization. Stat Methods Med Res. 2017 Oct;26(5):2333–55.

24. Nikpay M, Goel A, Won HH, Hall LM, Willenborg C, Kanoni S, Saleheen D, Kyriakou T, Nelson CP, Hopewell JC, Webb TR, Zeng L, Dehghan A, Alver M, Armasu SM, Auro K, Bjonnes A, Chasman DI, Chen S, Ford I, Franceschini N, Gieger C, Grace C, Gustafsson S, Huang J, Hwang SJ, Kim YK, Kleber ME, Lau KW, Lu X, Lu Y, Lyytikäinen LP, Mihailov E, Morrison AC, Pervjakova N, Qu L, Rose LM, Salfati E, Saxena R, Scholz M, Smith AV, Tikkanen E, Uitterlinden A, Yang X, Zhang W, Zhao W, de Andrade M, de Vries PS, van Zuydam NR, Anand SS, Bertram L, Beutner F, Dedoussis G, Frossard P, Gauguier D, Goodall AH, Gottesman O, Haber M, Han BG, Huang J, Jalilzadeh S, Kessler T, König IR, Lannfelt L, Lieb W, Lind L, Lindgren CM, Lokki ML, Magnusson PK, Mallick NH, Mehra N, Meitinger T, Memon F ur R, Morris AP, Nieminen MS, Pedersen NL, Peters A, Rallidis LS, Rasheed A, Samuel M, Shah SH, Sinisalo J, Stirrups KE, Trompet S, Wang L, Zaman KS, Ardissino D, Boerwinkle E, Borecki IB, Bottinger EP, Buring JE, Chambers JC, Collins R, Cupples LA, Danesh J, Demuth I, Elosua R, Epstein SE, Esko T, Feitosa MF, Franco OH, Franzosi MG, Granger CB, Gu D, Gudnason V, Hall AS, Hamsten A, Harris TB, Hazen SL, Hengstenberg C, Hofman A, Ingelsson E, Iribarren C, Jukema JW, Karhunen PJ, Kim BJ, Kooner JS, Kullo IJ, Lehtimäki T, Loos RJF, Melander O, Metspalu A, März W, Palmer CN, Perola M, Quertermous T, Rader DJ, Ridker PM, Ripatti S, Roberts R, Salomaa V, Sanghera DK, Schwartz SM, Seedorf U, Stewart AF, Stott DJ, Thiery J, Zalloua PA, O’Donnell CJ, Reilly MP, Assimes TL, Thompson JR, Erdmann J, Clarke R, Watkins H, Kathiresan S, McPherson R, Deloukas P, Schunkert H, Samani NJ, Farrall M, the CARDIoGRAMplusC4D Consortium. A comprehensive 1000 Genomes–based genome-wide association meta-analysis of coronary artery disease. Nat Genet. 2015 Oct;47(10):1121–30.

25. Malik R, Chauhan G, Traylor M, Sargurupremraj M, Okada Y, Mishra A, Rutten-Jacobs L, Giese AK, van der Laan SW, Gretarsdottir S, Anderson CD, Chong M, Adams HHH, Ago T, Almgren P, Amouyel P, Ay H, Bartz TM, Benavente OR, Bevan S, Boncoraglio GB, Brown RD, Butterworth AS, Carrera C, Carty CL, Chasman DI, Chen WM, Cole JW, Correa A, Cotlarciuc I, Cruchaga C, Danesh J, de Bakker PIW, DeStefano AL, den Hoed M, Duan Q, Engelter ST, Falcone GJ, Gottesman RF, Grewal RP, Gudnason V, Gustafsson S, Haessler J, Harris TB, Hassan A, Havulinna AS, Heckbert SR, Holliday EG, Howard G, Hsu FC, Hyacinth HI, Ikram MA, Ingelsson E, Irvin MR, Jian X, Jiménez-Conde J, Johnson JA, Jukema JW, Kanai M, Keene KL, Kissela BM, Kleindorfer DO, Kooperberg C, Kubo M, Lange LA, Langefeld CD, Langenberg C, Launer LJ, Lee JM, Lemmens R, Leys D, Lewis CM, Lin WY, Lindgren AG, Lorentzen E, Magnusson PK, Maguire J, Manichaikul A, McArdle PF, Meschia JF, Mitchell BD, Mosley TH, Nalls MA, Ninomiya T, O’Donnell MJ, Psaty BM, Pulit SL, Rannikmäe K, Reiner AP, Rexrode KM, Rice K, Rich SS, Ridker PM, Rost NS, Rothwell PM, Rotter JI, Rundek T, Sacco RL, Sakaue S, Sale MM, Salomaa V, Sapkota BR, Schmidt R, Schmidt CO, Schminke U, Sharma P, Slowik A, Sudlow CLM, Tanislav C, Tatlisumak T, Taylor KD, Thijs VNS, Thorleifsson G, Thorsteinsdottir U, Tiedt S, Trompet S, Tzourio C, van Duijn CM, Walters M, Wareham NJ, Wassertheil-Smoller S, Wilson JG, Wiggins KL, Yang Q, Yusuf S, AFGen Consortium, Cohorts for Heart and Aging Research in Genomic Epidemiology (CHARGE) Consortium, International Genomics of Blood Pressure (iGEN-BP) Consortium, INVENT Consortium, STARNET, Bis JC, Pastinen T, Ruusalepp A, Schadt EE, Koplev S, Björkegren JLM, Codoni V, Civelek M, Smith NL, Trégouët DA, Christophersen IE, Roselli C, Lubitz SA, Ellinor PT, Tai ES, Kooner JS, Kato N, He J, van der Harst P, Elliott P, Chambers JC, Takeuchi F, Johnson AD, BioBank Japan Cooperative Hospital Group, COMPASS Consortium, EPIC-CVD Consortium, EPIC-InterAct Consortium, International Stroke Genetics Consortium (ISGC), METASTROKE Consortium, Neurology Working Group of the CHARGE Consortium, NINDS Stroke Genetics Network (SiGN), UK Young Lacunar DNA Study, MEGASTROKE Consortium, Sanghera DK, Melander O, Jern C, Strbian D, Fernandez-Cadenas I, Longstreth WT, Rolfs A, Hata J, Woo D, Rosand J, Pare G, Hopewell JC, Saleheen D, Stefansson K, Worrall BB, Kittner SJ, Seshadri S, Fornage M, Markus HS, Howson JMM, Kamatani Y, Debette S, Dichgans M. Multiancestry genome-wide association study of 520,000 subjects identifies 32 loci associated with stroke and stroke subtypes. Nat Genet. 2018 Apr;50(4):524–37.

26. Thomas DC, Lawlor DA, Thompson JR. Re: Estimation of bias in nongenetic observational studies using “Mendelian triangulation” by Bautista et al. Ann Epidemiol. 2007 Jul;17(7):511–3.

27. Vaucher J, Keating BJ, Lasserre AM, Gan W, Lyall DM, Ward J, Smith DJ, Pell JP, Sattar N, Paré G, Holmes MV. Cannabis use and risk of schizophrenia: a Mendelian randomization study. Mol Psychiatry. 2018 May;23(5):1287–92.

28. Hemani G, Tilling K, Smith GD. Orienting the causal relationship between imprecisely measured traits using GWAS summary data. PLOS Genetics. 2017 Nov 17;13(11):e1007081.

29. Staiger D, Stock JH. Instrumental Variables Regression with Weak Instruments. Econometrica. 1997;65(3):557–86.

30. Bowden J, Davey Smith G, Burgess S. Mendelian randomization with invalid instruments: effect estimation and bias detection through Egger regression. Int J Epidemiol. 2015 Apr;44(2):512– 25.

31. Bowden J, Del Greco M F, Minelli C, Davey Smith G, Sheehan NA, Thompson JR. Assessing the suitability of summary data for two-sample Mendelian randomization analyses using MR-Egger regression: the role of the I2 statistic. Int J Epidemiol. 2016 Dec 1;45(6):1961–74.

32. Bowden J, Davey Smith G, Haycock PC, Burgess S. Consistent Estimation in Mendelian Randomization with Some Invalid Instruments Using a Weighted Median Estimator. Genet Epidemiol. 2016 May;40(4):304–14.

33. Rabin RA, George TP. A review of co-morbid tobacco and cannabis use disorders: possible mechanisms to explain high rates of co-use. Am J Addict. 2015 Mar;24(2):105–16.

34. Stringer S, Minică CC, Verweij KJH, Mbarek H, Bernard M, Derringer J, van Eijk KR, Isen JD, Loukola A, Maciejewski DF, Mihailov E, van der Most PJ, Sánchez-Mora C, Roos L, Sherva R, Walters R, Ware JJ, Abdellaoui A, Bigdeli TB, Branje SJT, Brown SA, Bruinenberg M, Casas M, Esko T, Garcia-Martinez I, Gordon SD, Harris JM, Hartman CA, Henders AK, Heath AC, Hickie IB, Hickman M, Hopfer CJ, Hottenga JJ, Huizink AC, Irons DE, Kahn RS, Korhonen T, Kranzler HR, Krauter K, van Lier P a. C, Lubke GH, Madden P a. F, Mägi R, McGue MK, Medland SE, Meeus WHJ, Miller MB, Montgomery GW, Nivard MG, Nolte IM, Oldehinkel AJ, Pausova Z, Qaiser B, Quaye L, Ramos-Quiroga JA, Richarte V, Rose RJ, Shin J, Stallings MC, Stiby AI, Wall TL, Wright MJ, Koot HM, Paus T, Hewitt JK, Ribasés M, Kaprio J, Boks MP, Snieder H, Spector T, Munafò MR, Metspalu A, Gelernter J, Boomsma DI, Iacono WG, Martin NG, Gillespie NA, Derks EM, Vink JM. Genome-wide association study of lifetime cannabis use based on a large meta-analytic sample of 32 330 subjects from the International Cannabis Consortium. Transl Psychiatry. 2016 Mar 29;6:e769.

35. Liu M, Jiang Y, Wedow R, Li Y, Brazel DM, Chen F, Datta G, Davila-Velderrain J, McGuire D, Tian C, Zhan X, Choquet H, Docherty AR, Faul JD, Foerster JR, Fritsche LG, Gabrielsen ME, Gordon SD, Haessler J, Hottenga JJ, Huang H, Jang SK, Jansen PR, Ling Y, Mägi R, Matoba N, McMahon G, Mulas A, Orrù V, Palviainen T, Pandit A, Reginsson GW, Skogholt AH, Smith JA, Taylor AE, Turman C, Willemsen G, Young H, Young KA, Zajac GJM, Zhao W, Zhou W, Bjornsdottir G, Boardman JD, Boehnke M, Boomsma DI, Chen C, Cucca F, Davies GE, Eaton CB, Ehringer MA, Esko T, Fiorillo E, Gillespie NA, Gudbjartsson DF, Haller T, Harris KM, Heath AC, Hewitt JK, Hickie IB, Hokanson JE, Hopfer CJ, Hunter DJ, Iacono WG, Johnson EO, Kamatani Y, Kardia SLR, Keller MC, Kellis M, Kooperberg C, Kraft P, Krauter KS, Laakso M, Lind PA, Loukola A, Lutz SM, Madden PAF, Martin NG, McGue M, McQueen MB, Medland SE, Metspalu A, Mohlke KL, Nielsen JB, Okada Y, Peters U, Polderman TJC, Posthuma D, Reiner AP, Rice JP, Rimm E, Rose RJ, Runarsdottir V, Stallings MC, Stančáková A, Stefansson H, Thai KK, Tindle HA, Tyrfingsson T, Wall TL, Weir DR, Weisner C, Whitfield JB, Winsvold BS, Yin J, Zuccolo L, Bierut LJ, Hveem K, Lee JJ, Munafo MR, Saccone NL, Willer CJ, Cornelis MC, David SP, Hinds D, Jorgenson E, Kaprio J, Stitzel JA, Stefansson K, Thorgeirsson TE, Abecasis G, Liu DJ, Vrieze S. Association studies of up to 1.2 million individuals yield new insights into the genetic etiology of tobacco and alcohol use. Nat Genet. 2019 Feb;51(2):237–44.

36. Burgess S, Davey Smith G, Davies NM, Dudbridge F, Gill D, Glymour MM, Hartwig FP, Holmes MV, Minelli C, Relton CL, Theodoratou E. Guidelines for performing Mendelian randomization investigations. Wellcome Open Res. 2020 Apr 28;4:186.

37. Johnson EC, Demontis D, Thorgeirsson TE, Walters RK, Polimanti R, Hatoum AS, Sanchez-Roige S, Paul SE, Wendt FR, Clarke TK, Lai D, Reginsson GW, Zhou H, He J, Baranger DAA, Gudbjartsson DF, Wedow R, Adkins DE, Adkins AE, Alexander J, Bacanu SA, Bigdeli TB, Boden J, Brown SA, Bucholz KK, Bybjerg-Grauholm J, Corley RP, Degenhardt L, Dick DM, Domingue BW, Fox L, Goate AM, Gordon SD, Hack LM, Hancock DB, Hartz SM, Hickie IB, Hougaard DM, Krauter K, Lind PA, McClintick JN, McQueen MB, Meyers JL, Montgomery GW, Mors O, Mortensen PB, Nordentoft M, Pearson JF, Peterson RE, Reynolds MD, Rice JP, Runarsdottir V, Saccone NL, Sherva R, Silberg JL, Tarter RE, Tyrfingsson T, Wall TL, Webb BT, Werge T, Wetherill L, Wright MJ, Zellers S, Adams MJ, Bierut LJ, Boardman JD, Copeland WE, Farrer LA, Foroud TM, Gillespie NA, Grucza RA, Harris KM, Heath AC, Hesselbrock V, Hewitt JK, Hopfer CJ, Horwood J, Iacono WG, Johnson EO, Kendler KS, Kennedy MA, Kranzler HR, Madden PAF, Maes HH, Maher BS, Martin NG, McGue M, McIntosh AM, Medland SE, Nelson EC, Porjesz B, Riley BP, Stallings MC, Vanyukov MM, Vrieze S, Davis LK, Bogdan R, Gelernter J, Edenberg HJ, Stefansson K, Børglum AD, Agrawal A. A large-scale genome-wide association study meta-analysis of cannabis use disorder. Lancet Psychiatry. 2020 Dec;7(12):1032–45.

38. Watanabe AH, Navaravong L, Sirilak T, Prasitwarachot R, Nathisuwan S, Page RL, Chaiyakunapruk N. A systematic review and meta-analysis of randomized controlled trials of cardiovascular toxicity of medical cannabinoids. J Am Pharm Assoc (2003). 2021 Oct;61(5):e1– 13.

39. Winhusen T, Theobald J, Kaelber DC, Lewis D. The association between regular cannabis use, with and without tobacco co-use, and adverse cardiovascular outcomes: cannabis may have a greater impact in non-tobacco smokers. Am J Drug Alcohol Abuse. 2020 Jul 3;46(4):454–61.

40. Richards JR, Bing ML, Moulin AK, Elder JW, Rominski RT, Summers PJ, Laurin EG. Cannabis use and acute coronary syndrome. Clinical Toxicology. 2019 Oct 3;57(10):831–41.

41. Chami T, Kim CH. Cannabis Abuse and Elevated Risk of Myocardial Infarction in the Young: A Population-Based Study. Mayo Clin Proc. 2019 Aug;94(8):1647–9.

42. DeFilippis EM, Bajaj NS, Singh A, Malloy R, Givertz MM, Blankstein R, Bhatt DL, Vaduganathan M. Marijuana Use in Patients with Cardiovascular Disease: Current Knowledge and Gaps. J Am Coll Cardiol. 2020 Jan 28;75(3):320–32.

43. Latif Z, Garg N. The Impact of Marijuana on the Cardiovascular System: A Review of the Most Common Cardiovascular Events Associated with Marijuana Use. J Clin Med. 2020 Jun 19;9(6):1925.

44. Causal Inference: What If (the book) | Miguel Hernan’s Faculty Website | Harvard T.H. Chan School of Public Health [Internet]. [cited 2022 Feb 4]. Available from: https://www.hsph.harvard.edu/miguel-hernan/causal-inference-book/

45. Mounier N, Kutalik Z. Correction for sample overlap, winner’s curse and weak instrument bias in two-sample Mendelian Randomization [Internet]. bioRxiv; 2021 [cited 2022 Nov 29]. p. 2021.03.26.437168. Available from: https://www.biorxiv.org/content/10.1101/2021.03.26.437168v1

46. Burgess S, Davies NM, Thompson SG. Bias due to participant overlap in two-sample Mendelian randomization. Genet Epidemiol. 2016 Nov;40(7):597–608.

