## Supplementary_Material for "Cannabis use and atherosclerotic cardiovascular disease: a Mendelian randomization study"

### Supplementary Material

Roxane de La Harpe et al.

### Supplementary Methodology

#### Transformation of MR causal estimates from a per-1-log unit increase in ever use of cannabis to users vs. non-users

It is based on Ross et al's equations[1] and descriptions below come from Vaucher et al[2]:

Given:

(A) Prevalence of global lifetime cannabis use = 0.272 for European population.[3]

(B) Prevalence of CAD and IS in non-users of cannabis from an observational cohort.

(C) Odds ratio for CAD and IS associated with genetically determined cannabis use (users vs. non users) at the population level

(D) Genetic association with CAD and IS as function of genetic association with use of cannabis (where causal genetic effects are expressed as Log OR per allele for both outcome and use of cannabis)

The following can be calculated:

(E) Calculated population prevalence of CAD and IS: **(A × B × C) + (1 - A) × B**

(F) Estimated prevalence of CAD and IS in individuals with a theoretical increase in risk of use of cannabis of *e* = 2.72 fold: **(A × exp(1) × B × C) + (1 - A × exp(1)) × B**

(G) Estimated odds ratio for CAD and IS per *e* = 2.72 fold increase in risk of use of cannabis: **F/E**

It results that:

**(H)** $$\exp(\mathbf{D}) = \mathbf{G} = \frac{F}{E} = \frac{(\mathbf{A} \times \exp(1) \times \mathbf{B} \times \mathbf{C}) + (1 - \mathbf{A} \times \exp(1)) \times \mathbf{B}}{(\mathbf{A} \times \mathbf{B} \times \mathbf{C}) + (1 - \mathbf{A}) \times \mathbf{B}}$$

As C is the only unknown variable, the association between genetically determined use of cannabis (cannabis users vs. non-users) and risk of ASCVD (expressed as an odds ratio) at the population level can be calculated using algebraic transformations and (H) can be simplified into:

**(I)** $$\mathbf{C} = 1 + \frac{(1 - \exp(\mathbf{D}))}{(\exp(\mathbf{D}) - \exp(1)) \times \mathbf{A})}$$

#### Variance and F-statistic calculation for each SNP and cumulatively

The proportion of variance (conceptually similar to the $R^2$) in use of cannabis was computed for each SNP based on the formula provided by Shim et al.[4]

$$R^2 = \frac{2\beta^2 \text{ x MAF x } (1 - \text{MAF})}{2\beta^2 \text{ x MAF x } (1 - \text{MAF}) + (\text{se}(\beta))^2 \text{ x } 2N \text{ x MAF x } (1 - \text{MAF})}$$

with β, effect size (β coefficient) for a given SNP; MAF, minor allele frequency; se(β) standard error of effect size and *N*, sample size.

Total proportion of variance explained for the association between the SNPs and the use of cannabis result in cumulative effect of each SNP.

The F-statistics was then calculated as follow, according to Pierce et al[5] :

$$F = \frac{R^2(n-1-k)}{(1-R^2)k}$$

with, n, sample size; k, number of instrument variables used

**Association between cannabis use and ASCVD in CoLaus/PsyCoLaus cohort study**

The detailed description of the recruitment of the PsyCoLaus study and the follow-up procedures have been described previously[6]. Briefly, this cohort study is a population-based cohort exploring the biological, genetic and environmental determinants of ASCVD. A non-stratified, representative sample of the population of Lausanne (Switzerland) was recruited between 2003 and 2006 based on the following inclusion criteria: (1) age 35–75 years and (2) willingness to participate. The third follow-up occurred 15 years after the baseline survey.

ASCVD, comprised non-fatal acute myocardial infarction (AMI), symptomatic coronary artery disease with greater than 50% stenosis treated by percutaneous coronary intervention or coronary artery bypass graft (CHD), and fatal and non-fatal ischaemic stroke (including transient ischaemic attack) were comprehensively collected and independently adjudicated.

Participants were invited to attend the outpatient clinic at Lausanne University Hospital for a specific interview conducted by psychologists with questions relative to drug use, abuse and dependence, constructed the PsychoLaus cohort.

6’770 participants were involved at baseline and 5’128 took part to the PsyCoLaus Cohort. We excluded participant with missing data at baseline (n=434), previous ASCVD (n=93) and loss of follow-up (n=262).

Cox proportional hazard regressions with time of follow-up as the underlying time variable were used to test associations between cannabis use and incident ASCVD or stroke or coronary artery disease only. Co-variables were demographic variables (age and sex) and risk factors for ASCVD (body mass index, hypertension arterial and diabetes mellitus). Outcomes were censored if a participant was lost to follow-up, died from a non-cardiovascular cause, or if the end of available follow-up was reached.

**Genome-wide association study meta-analysis of cannabis use disorder**

We used summary-level data from the publicly available GWAS of Johnson et al, comprising 374’287 European participants, which explained phenotype defined as a lifetime diagnosis of cannabis abuse or dependence (according to DSM-IV or ICD_10). The GWAS was performed of 20 samples (18 from the Psychiatric Genomic Consortium Substance Use Disorder working group, on from iPSYCH and one from decoDE, More information about recruitment, measurement instruments and phrasing of the questions about lifetime cannabis use can be found in the supplementary material of Johnson et al.[7]We selected SNPs with p-value that were associated with cannabis use disorder (p-value<$5x10^{-5}$)

and in low linkage-disequilibrium with other SNPs ($R^2 < 0.001$) within a clumping distance of 10,000 kb. Palindromic SNPs with intermediate minor allele frequency were removed because we cannot assumed that all alleles were correctly reported in the positive strand in cannabis use disorder GWAS. A more restricted threshold with p-value $<5x10^{-8}$ was initially computed but only 2 SNPs remained, which was not sufficient for two-sample MR analysis (see Supplementary Table 10).

### Supplementary Tables

**Supplementary Table 1.** Studies included in the cannabis-GWAS used in our analysis

| Study | Country | *N total* | Reference/PMID |
|---|---|---|---|
| *International Cannabis Consortium* | | | |
| ALSPAC | UK | 2976 | 22507743 |
| BLTS | Australia | 721 | 23187020 |
| CADD | US | 853 | Not published |
| EGCUT1 | Estonia | 2765 | 15133739 |
| EGCUT2 | Estonia | 970 | 15133739 |
| FinnTwin | Finland | 1029 | 23298696 |
| HUVH | Spain | 981 | 25284319 |
| MCTFR | US | 6241 | 23363460 |
| NTR | Netherlands | 4653 | 20477721 |
| QIMR | Australia | 6778 | 17988414 |
| TRAILS | Netherlands | 1226 | 18763693 |
| Utrecht | Netherlands | 1173 | 20925969 |
| Radar | Dutch | 338 | 25466800 |
| SYS | Canada | 551 | 25454417 |
| TwinsUK | UK | 2078 | twinsuk.ac.uk |
| Yale Penn African American | US | 2660 | 24166409 |
| Yale Penn European American | US | 1964 | 24166409 |
| *23andMe* | | | |
| 23andMe | - | 22683 | - |
| *UK-Biobank* | | | |
| UK-Biobank | UK | 126785 | 25826379 |

Overview of studies included in the GWAS from Pasman et al[8]. In orange, studies which were also found in CARDIoGRAMplusC4D GWAS. No similar studies were found in MEGASTROKE.

**Supplementary Table 2.** Summary of the 65 SNPs associated with use of cannabis (by decreasing p-value)

| SNP | EA | EAF | Beta | Standard error | p-value | Chromosome | Sample size | $R^2$ | F-Statistic |
|---|---|---|---|---|---|---|---|---|---|
| rs2875907 | A | .3524 | .0712 | .0086 | 9.381e-17 | 3 | 184765 | .0003708 | 68.51757 |
| rs9919557 | C | .386 | .0549 | .0085 | 9.935e-11 | 11 | 184765 | .0002257 | 41.70699 |
| rs10499 | A | .6513 | .0532 | .0087 | 1.134e-09 | 16 | 184765 | .0002023 | 37.38496 |
| rs9773390 | C | .0673 | .1714 | .0294 | 5.659e-09 | 8 | 57980 | .0005859 | 33.96819 |
| rs17761723 | T | .3462 | .0473 | .0085 | 3.236e-08 | 17 | 184765 | .0001676 | 30.96076 |
| rs466765 | A | .2092 | .0565 | .0104 | 5.884e-08 | 6 | 184765 | .0001597 | 29.50943 |
| rs1154693 | G | .8538 | .063 | .0117 | 6.921e-08 | 3 | 184765 | .0001569 | 28.98953 |
| rs12373990 | T | .1171 | .0667 | .0128 | 1.989e-07 | 22 | 184765 | .0001469 | 27.14988 |
| rs6827295 | G | .7139 | .0467 | .0091 | 2.490e-07 | 4 | 184765 | .0001425 | 26.33232 |
| rs12211611 | G | .8084 | .0538 | .0104 | 2.552e-07 | 6 | 184765 | .0001448 | 26.75685 |
| rs1066339 | A | .1683 | .1472 | .0286 | 2.719e-07 | 6 | 57980 | .0004567 | 26.478 |
| rs9972422 | G | .2907 | .0462 | .009 | 2.754e-07 | 15 | 184765 | .0001426 | 26.34735 |
| rs437021 | C | .5408 | .0422 | .0082 | 2.770e-07 | 1 | 184765 | .0001433 | 26.48103 |
| rs576076 | A | .2534 | .0477 | .0094 | 4.431e-07 | 11 | 184765 | .0001393 | 25.74664 |
| rs114212469 | T | .0207 | .1578 | .0317 | 6.230e-07 | 4 | 149468 | .0001658 | 24.77556 |
| rs1808579 | T | .4792 | .0408 | .0082 | 6.804e-07 | 18 | 184765 | .000134 | 24.75338 |
| rs9435794 | C | .289 | .046 | .0094 | 9.198e-07 | 1 | 184765 | .0001296 | 23.94439 |
| rs7871607 | G | .0117 | .1982 | .0404 | 9.205e-07 | 9 | 149468 | .000161 | 24.06438 |
| rs205723 | A | .4141 | .041 | .0084 | 1.029e-06 | 7 | 184765 | .0001289 | 23.82063 |
| rs60369116 | G | .9754 | .1428 | .0295 | 1.241e-06 | 7 | 149468 | .0001567 | 23.42849 |
| rs11902472 | A | .6168 | .0405 | .0084 | 1.312e-06 | 2 | 184765 | .0001258 | 23.24325 |

| | | | | | | | | | |
|---|---|---|---|---|---|---|---|---|---|
| rs4377758 | G | .0561 | .0943 | .0195 | 1.339e-06 | 6 | 184765 | .0001266 | 23.38294 |
| rs79294243 | C | .0441 | .0989 | .0206 | 1.639e-06 | 2 | 184765 | .0001247 | 23.04644 |
| rs10849982 | G | .8283 | .052 | .0109 | 1.762e-06 | 12 | 184765 | .0001232 | 22.75623 |
| rs2059730 | G | .6686 | .0419 | .0088 | 1.854e-06 | 2 | 184765 | .0001227 | 22.6678 |
| rs11209802 | C | .6795 | .042 | .0088 | 1.957e-06 | 1 | 184765 | .0001233 | 22.77612 |
| rs146752096 | T | .0862 | .0691 | .0145 | 1.960e-06 | 2 | 184765 | .0001229 | 22.70736 |
| rs1012534 | A | .5662 | .0388 | .0082 | 2.251e-06 | 6 | 184765 | .0001212 | 22.38634 |
| rs1885331 | T | .7508 | .045 | .0095 | 2.333e-06 | 6 | 184765 | .0001214 | 22.43495 |
| rs2305758 | T | .2802 | .0425 | .009 | 2.699e-06 | 19 | 184765 | .0001207 | 22.29669 |
| rs78698099 | G | .9496 | .1008 | .0215 | 2.779e-06 | 16 | 149468 | .000147 | 21.9776 |
| rs13123620 | A | .5914 | .0387 | .0083 | 3.196e-06 | 4 | 184765 | .0001177 | 21.73775 |
| rs79777905 | G | .9824 | .1716 | .0369 | 3.315e-06 | 8 | 149468 | .0001447 | 21.62315 |
| rs9855698 | G | .1398 | .0564 | .0122 | 3.728e-06 | 3 | 184765 | .0001157 | 21.3692 |
| rs1587858 | C | .301 | .042 | .0091 | 3.786e-06 | 5 | 184765 | .0001153 | 21.29932 |
| rs1503510 | C | .6503 | .0397 | .0086 | 3.939e-06 | 5 | 184765 | .0001153 | 21.30758 |
| rs9655332 | T | .4229 | .0668 | .0145 | 3.953e-06 | 7 | 57980 | .0003659 | 21.21573 |
| rs701802 | G | .6396 | .0412 | .009 | 4.135e-06 | 10 | 184765 | .0001134 | 20.95367 |
| rs7513688 | G | .6418 | .0394 | .0086 | 4.480e-06 | 1 | 184765 | .0001136 | 20.9868 |
| rs2086512 | A | .1093 | .0596 | .013 | 4.552e-06 | 6 | 184765 | .0001137 | 21.01631 |
| rs73262787 | G | .9468 | .0861 | .0188 | 4.626e-06 | 12 | 184765 | .0001135 | 20.97207 |
| rs6948053 | G | .056 | .0856 | .0187 | 4.660e-06 | 7 | 184765 | .0001134 | 20.9515 |
| rs830147 | G | .9471 | .1703 | .0372 | 4.725e-06 | 19 | 57980 | .0003613 | 20.95012 |
| rs4837004 | C | .3402 | .0394 | .0086 | 4.787e-06 | 9 | 184765 | .0001136 | 20.9868 |

| | | | | | | | | | |
|---|---|---|---|---|---|---|---|---|---|
| rs4147187 | T | .021 | .1434 | .0314 | 4.952e-06 | 9 | 149468 | .0001395 | 20.85347 |
| rs149434117 | T | .9829 | .1642 | .036 | 4.962e-06 | 3 | 149468 | .0001392 | 20.80084 |
| rs62638743 | A | .0296 | .1157 | .0254 | 5.096e-06 | 19 | 184765 | .0001123 | 20.74677 |
| rs12949052 | A | .9258 | .073 | .016 | 5.103e-06 | 17 | 184765 | .0001127 | 20.81406 |
| rs6047198 | C | .7527 | .0433 | .0095 | 5.409e-06 | 20 | 184765 | .0001124 | 20.77207 |
| rs11724871 | C | .4671 | .0378 | .0083 | 5.766e-06 | 4 | 184765 | .0001122 | 20.73856 |
| rs7969834 | G | .273 | .0413 | .0092 | 6.578e-06 | 12 | 184765 | .0001091 | 20.15009 |
| rs10012797 | A | .9167 | .0684 | .0152 | 6.631e-06 | 4 | 184765 | .0001096 | 20.24778 |
| rs143529057 | C | .989 | .2036 | .0453 | 6.891e-06 | 5 | 149468 | .0001351 | 20.19763 |
| rs17294232 | A | .5541 | .0377 | .0084 | 7.105e-06 | 2 | 184765 | .000109 | 20.1408 |
| rs61997596 | A | .1862 | .0484 | .0108 | 7.111e-06 | 14 | 184765 | .0001087 | 20.08149 |
| rs79563551 | C | .9778 | .1427 | .0319 | 7.499e-06 | 4 | 149468 | .0001339 | 20.00822 |
| rs17481131 | T | .7925 | .0452 | .0101 | 8.062e-06 | 12 | 184765 | .0001084 | 20.02567 |
| rs139621111 | C | .1961 | .0877 | .0197 | 8.150e-06 | 7 | 57980 | .0003417 | 19.81154 |
| rs7670670 | T | .2236 | .0444 | .0099 | 8.177e-06 | 4 | 184765 | .0001089 | 20.11168 |
| rs80144387 | T | .0699 | .0857 | .0193 | 8.757e-06 | 2 | 162082 | .0001216 | 19.71488 |
| rs2049824 | C | .4674 | .0365 | .0082 | 9.076e-06 | 8 | 184765 | .0001072 | 19.81123 |
| rs10849767 | T | .3344 | .0397 | .009 | 9.384e-06 | 12 | 184765 | .0001053 | 19.45585 |
| rs72798040 | T | .9009 | .0624 | .0141 | 9.397e-06 | 16 | 184765 | .000106 | 19.58326 |
| rs4445597 | T | .1045 | .0603 | .0136 | 9.856e-06 | 11 | 184765 | .0001064 | 19.6567 |
| rs7758880 | C | .7075 | .036 | .0092 | .00008703 | 6 | 184765 | .0000829 | 15.31064 |

We computed for each SNP the variance ($R^2$) and the F-statistic, as explained in Supplmentary Methods.
None of the 65 SNPs were in linkage disequilibrium based on SNP Annotation and Proxy Search (SNAP, Broad Institute, MA, US https://data.broadinstitute.org/mpg/snpsnap/ ) with a $R^2$ threshold < 0.2.

In red, SNP excluded because insufficient significant p-value. In blue, SNP not found in CAD-GWAS summary statistics
Chr, chromosome; EAF, effect allele frequency; SE, standard error. Beta coefficient corresponds to log odds of ever use of cannabis. SNP, single-nucleotide polymorphism; EA, effect allele; EAF, effect allele frequency; Beta, the per-allele effect on cannabis use from the meta-analysis; Standard error of Beta; p-value is for the genetic association in the GWAS.; Sample size, depending on in how many samples this SNP was present.

**Supplementary Table 3.** Summary of the observational studies included in the analysis of cannabis use and risk of ASCVD, presented by specific outcome and type of study design

| Type of study (country, years of recruitment) | Years of follow-up | Cases* | Controls | Exposure (as reported in the text) | Outcome (source) | Adjustments | Reference (PMID) |
|---|---|---|---|---|---|---|---|
| **Coronary artery disease including acute myocardial infarction** | | | | | | | |
| Cohort prospective (USA; 1985-1986) | 27 | 104 | 5009 | Cumulative marijuana use (never vs. min 0.5 marijuana-years) | CHD (telephone interviews, examinations, medical records) | Age, gender, race, education, family history of ASCVD, physical activity, BMI, HTA, DM, dyslipidemia, depression, smoking, cumulative alcohol use, cumulative use of other illicit drugs | Reis et al[9] 2017 (28207342) |
| Cohort prospective (Swiss, 2003-2006) | 13 | 227 | 4112 | Cannabis use (never vs. abuse or dependence) | AMI and CHD (Electronic records with ICD codes) | Age, sex, others substances abuse , smoking, depression, stress disorders, psychotic disorders, BMI | See Supplemtentary Methods |
| Cohort retrospective (USA, 2011-2016) | 3 | 60940 | 10'774'178 | History of cannabis abuse (ever v. never) | AMI (from multi-institutional database) | Age, sex, hypertension, coronary artery disease, diabetes, and other substance abuse | Chami et al[10], 2019 (31378243) |
| A case - matched cohort study (USA, 2016-2018) | 2 | 84 | 465 | Cannabis use (never vs. use, abuse or dependence) | CHD (Electronic ICD-10$^{th}$ codes) | BMI, smoking, households, others substances abuse, any chronic health condition, any acute health events | Philips et al[11], 2022 (35279458) |

| | | | | | | | |
|---|---|---|---|---|---|---|---|
| Case-control study (USA, 2010-2014) | - | 1694 | 9'465’255 | Cannabis use (never vs. current cannabis use disorder) | AMI (Electronic records with ICD-codes) | Demographics, medical risk factors, and others substances abuse | Patel et al[12] 2020 (31611137**)** |
| Case-control study (USA 2016-2017) | - | 201 | 1219 | Marijuana use (never vs. ever) | CHD according to CT angiogram | Age, DM, HTA | Burt et al[13] 2020 (31995626) |
| **Ischemic stroke and transient ischemic attack** | | | | | | | |
| Cohort prospective (Sweden, 1949-1951) | 26 | 1037 | 44044 | Cannabis use (never vs. 1 to 10 times) | IS and TIA (Electronic records with ICD-8th and -9th codes) | Age, BMI, migraine, DM, family history of ASCVD, HTA, cardiorespiratory fitness, childhood socioeconomic position, short schooling, smoking, alcohol consumption | Falksted et al[14], 2017 (28028147) |
| Cohort prospective (USA; 1985-1986) | 27 | 62 | 5051 | Cumulative marijuana use (never vs. min 0.5 marijuana-years) | IS and TIA (telephone interviews, examinations, medical records) | Age, gender, race, education, family history of ASCVD, physical activity, BMI, HTA, DM, dyslipidemia, depression, smoking, cumulative alcohol use, cumulative use of other illicit drugs | Reis et al[9] 2017 (2807342) |
| Cohort prospective (Swiss, 2003-2006) | 13 | 79 | 4260 | Cannabis use (never vs. abuse or dependence) | IS and TIA (Electronic records with ICD codes) | Age, sex, others substances abuse, smoking, depression, stress disorders, psychotic disorders, BMI | See Supplemtentary Methods |
| Case-control study (USA 2004-2011) | - | 478’650 | 118’180’969 | Marijuana use (never vs. cannabis use disorders) | IS and vasospam (Electronic records with ICD-9th codes) | Age, sex, race, payer status, Charlson’s comorbidity index, substances abuse | Rumalla et al[15], 2016 (26874461) |

| | | | | | | | |
|---|---|---|---|---|---|---|---|
| Cross-sectional study (USA, 2000-2003) | - | 998 | 811’249 | Cannabis use (ever vs. cannabis abuse or dependence) | IS (Electronic records with ICD-9th codes) | Others addictive substances, cardiovascular comorbidities | Westover et al[16], 2007 (17404126) |
| Cross-sectional study (USA, 2007-2011) | - | 105957 | 3’949’000 | Cannabis use (ever vs. cannabis abuse or dependence) | IS (Electronic records with ICD-9th codes) | Age, gender, race, residential income, insurance, residential region, pain, and number of comorbidities | Vin Raviv et al[17],2017 (27891823) |
| **ASCVD mortality or all ASCVD** | | | | | | | |
| Cohort prospective (USA, 2005-2014) | 6 | 39 | 14818 | Cannabis use (never vs. ever) | ASCVD mortality including heart disease and cerebrovascular disease (Electronic records with ICD codes) | Age, sex, race/ethnicity, education, family income level, smoking, alcohol intake, physical activity, diet, BMI | Sun et al[18] 2020 (33220757) |
| Cohort prospective (USA; 1985-1986) | 27 | 215 | 4898 | Cumulative marijuana use (never vs. min 0.5 marijuana-years) | ASCVD mortality (telephone interviews, examinations, medical records) | Age, gender, race, education, family history of ASCVD, physical activity, BMI, HTA, DM, dyslipidemia, depression, smoking, cumulative alcohol use, cumulative use of other illicit drugs | Reis et al[9] 2017 (28207342) |
| Cohort prospective (Canada, 1989) | 20 | 63923 | 1’247’035 | Cannabis use (never vs. current or past history of cannabis use disorders) | ASCVD (Electronic records with ICD codes) | Age, gravidity, mental illness, smoking, comorbidity, socioeconomic deprivation, place of residence, time-period | Auger et al[19] 2020 (33208143) |

| | | | | | | | |
|---|---|---|---|---|---|---|---|
| Cohort prospective (Swiss, 2003-2006) | 12 | 299 | 4040 | Cannabis use (never vs. abuse or dependence) | ASCVD (Electronic records with ICD codes) | Age, sex, BMI, HTA, DM, smoking | See Supplemtentary Methods |

AMI, acute myocardial infarction ; CHD, coronary heart disease; BMI, body mass index; HTA, hypertension arterial; IS, acute ischemic stroke; TIA, transient ischemic attack.* cases involving participants with outcome event.

**Supplementary Table 4.** Summary of the observational studies excluded in the analysis of cannabis use and risk of ASCVD, presented by specific outcome and type of study

| **Type of study (country, years of recruitment)** | **Reason of exclusion** | **Years of follow-up** | **Exposure** | **Outcome (source)** | **Adjustments (results: RR, 95% CI)** | **Reference (PMID)** |
|---|---|---|---|---|---|---|
| **Coronary artery disease including acute myocardial infarction** | | | | | | |
| Cohort retrospective (USA,-) | Comparison group without never use | 7,5 | Cannabis use (non-frequent or never vs. frequent) | AMI (Electronic records with ICD-9$^{th}$ and -10$^{th}$ codes) | Other substance use disorders, time since the earliest observation of cannabis use, age, sex, race, ethnicity, median income, BMI **(OR, 1.25, 0.80 – 1.95)** | Winhusen et al 2019 (31743053) |
| **Ischemic stroke and transient ischemic attack** | | | | | | |
| Cohort retrospective (USA,-) | Comparison group without never use | 7,5 | Cannabis use (non-frequent or never vs. frequent) | AMI (Electronic records with ICD-9$^{th}$ and -10$^{th}$ codes) | Other substance use disorders, time since the earliest observation of cannabis use, age, sex, race, ethnicity, median income, BMI **(OR, 1.62, 1.05–2.50)** | Winhusen et al 2019 (31743053) |
| Cross-sectional study (Australia, 1999-2001) | Lifetime use of cannabis not available | - | Marijuana user (never vs. recent marijuana user) | Self-reported IS or TIA | Age, level of education, working status, smoking, HTA, DM, exercise frequency **(OR, 2.3, 1.1-4.5)** | Hemachandra et al 2016 (26558539) |
| Cross-sectional study (USA, 2009-2010) | Imprecise outcome | - | Cannabis use (ever vss. Cannabis use disorders) | Cerebrovascular accident | Age, sex, HTA, DM, hyperlipidemia, CAD, smoking, and alcohol use **(OR 1.24, 1.14-1.34)** | Kalla et al 2018 (29879084) |

| | | | | | | |
|---|---|---|---|---|---|---|
| Cross-sectional study (USA, 2016-2017) | Lifetime use of cannabis not available | - | Marijuana user (never vs. recent marijuana user) | Self-reported IS | Age, sex, race, education, marital status, BMI, physical activity, smoking, e-cigarette use, heavy drinking, DM<br>**(OR 1.82, 1.08–3.10)** | Parekh et al 2020 (31707926) |
| **ASCVD mortality or all ASCVD** | | | | | | |
| Cohort prospective (USA, 1989-1994) | Lifetime use of cannabis not available | 4 | Marijuana use (never vs. recent use [1 year before inclusion]) | ASCVD mortality (death certificates from state offices of vital records) | Age and sex<br>**(HR 1.9, 0.6−6.3)** | Mukamal et al, 2008 (18294478) |
| Cohort prospective (USA, 2000-2016) | Lifetime use of cannabis not available | 11 | Marijuana use (never vs. recent use [one week before inclusion]) | ASVCD mortality (Electronic records with ICD codes) | Age, sex, DM, HTA, peripheral vascular disease, smoking, HDL-C, triglycerides, revascularization, creatinine, medications at discharge, length of stay<br>**(HR 2.09, 1.25- 3.5)** | DeFilipis et al, 2018 (29535062) |
| Cohort prospective (USA, 1990-2010) | Comparison group without never use | 6 | Heavy cannabis users (occasional use or nonuse vs. daily or weekly use | ASCVD (Electronic record with ICD-9$^{th}$ codes) | Age, smoking, viral load, traditional cardiovascular risk factor<br>**(HR 2.16 1.04-4.51)** | Lorenz et al 2017 (28449059) |

AMI, acute myocardial infarction ; CHD, coronary heart disease; BMI, body mass index; HTA, hypertension arterial; IS, acute ischemic stroke; TIA, transient ischemic attack.

**Supplementary Table 5**. Power (two-sided α=0.05) for conventional Mendelian randomization analysis

| | Exposure | Actual N in outcome-GWAS | Proportion of cases in outcome-GWAS | Observational OR | R2 of instrument | N required for 80% power | Power at actual N |
|---|---|---|---|---|---|---|---|
| **For CAD** | **Cannabis use** | 184’305 | 0.330 | 1.23* | 0.01 | 76895 | 0.99 |
| **For IS** | **Cannabis use** | 440328 | 0.084 | 1.22* | 0.01 | 217837 | 0.98 |

Power calculation was based on the method developed by Brion et al.[13] https://shiny.cnsgenomics.com/mRnd/ * from observational meta-analysis for CAD and IS respectively (Figure 1).

**Supplementary Table 6.** Comparison of conventional MR, MR-Egger, MR-Egger adjusted for SIMEX and weighted median MR causal effect estimates of cannabis use on risk of coronary artery disease

| **Analysis** | **Causal effect estimate** | **95% CI** |
|---|---|---|
| **Conventional MR** | -0.03 | -0.09-0.03 |
| **MR-Egger** ($I^2$=0.18) | -0.01 | -0.13; 0.11 |
| **MR-Egger+SIMEX** | -0.03 | -0.13- 0.08 |
| **Weighted median MR** | -0.05 | -0.13; 0.03 |

Estimates are expressed as Log OR per-1-log unit increase in ever use of cannabis. Conventional MR was pooled effect across SNPs using fixed-effect with inverse variance weighted meta-analysis. As previously described by Bowden et al [20], it is noteworthy to mention that power to detect a causal effect using MR-Egger analysis is largely underpowered (as shown by the corresponding large confidence intervals) with the use of 63 SNPs. The adjusted MR-Egger regression estimate (derived by simulation extrapolation [SIMEX] to account for a potential violation of the NOME assumption) is the result of 100,000 simulations. $I^2$ quantifies weak instrument bias in the context of MR-Egger. *for significant p-value (p-value<0.05)

**Supplementary Table 7.** Comparison of conventional MR, MR-Egger, MR-Egger adjusted for SIMEX and weighted median MR causal effect estimates of cannabis use on risk of acute ischemic stroke

| **Analysis** | **Causal effect estimate** | **95% CI** |
|---|---|---|
| **Conventional MR** | 0.08 | -0.05-0.10 |
| **MR-Egger** ($I^2$=0.39) | 0.04 | -0.11-0.18 |
| **MR-Egger+SIMEX** | -0.01 | -0.15-0.12 |
| **Weighted median MR** | 0.02 | -0.07-0.11 |

Estimates are expressed as Log OR per-1-log unit increase in ever use of cannabis. Conventional MR was pooled effect across SNPs using fixed-effect with inverse variance weighted meta-analysis. As previously described by Bowden et al, it is noteworthy to mention that power to detect a causal effect using MR-Egger analysis is largely underpowered (as shown by the corresponding large confidence intervals) with the use of 64 SNPs. The adjusted MR-Egger regression estimate (derived by simulation extrapolation [SIMEX] to account for a potential violation of the NOME assumption) is the result of 100,000 simulations. $I^2$ quantifies weak instrument bias in the context of MR-Egger. *for significant p-value (p-value<0.05)

**Supplementary Table 8.** Comparison of conventional MR, MR-Egger and weighted median MR causal effect estimates of cannabis use on ASVCD restricted to 5 SNPs with p-value<$5x10^{-8}$

| | **Coronary artery disease** | | **Acute ischemic stroke** | |
|---|---|---|---|---|
| **Analysis** | **Causal effect estimate** | **95% CI** | **Causal effect estimate** | **95% CI** |
| **Conventional MR** | -0.13 | -0.26-0.01 | 0.03 | -0.12-0.17 |
| **MR-Egger** | -0.07 | -0.41-0.27 | -0.12 | -0.47-0.22 |
| **Weighted median MR** | -0.12 | -0.28-0.05 | -0.01 | -0.19-0.16 |

Estimates are expressed as Log OR per-1-log unit increase in ever use of cannabis. Conventional MR was pooled effect across SNPs using fixed-effect with inverse variance weighted meta-analysis. No evidence against the null hypothesis of no directional pleiotropy of the genetic markers using MR-Egger was found (P-value for pleiotropy=0.730 for CAD and = 0.346 for IS). *for significant p-value (p-value<0.05)

**Supplementary Table 9.** Comparison of conventional MR, MR-Egger and weighted median MR causal effect estimates of cannabis use on ASVCD with removing SNPs being palindromic with intermediate allele frequencies.

| | Coronary artery disease | | | | Acute ischemic stroke | | | |
|---|---|---|---|---|---|---|---|---|
| | 64 SNPs (p-value<$5x10^{-5}$) | | 4 SNPs (p-value<$5x10^{-8}$) | | 65 SNPs (p-value<$5x10^{-5}$) | | 4 SNPs (p-value<$5x10^{-8}$) | |
| **Analysis** | **Causal effect estimate** | **95% CI** | **Causal effect estimate** | **95% CI** | **Causal effect estimate** | **95% CI** | **Causal effect estimate** | **95% CI** |
| **Conventional MR** | -0.03 | -0.08-0.02 | -0.13 | -0.31-0.05 | 0.03 | -0.04-0.10 | 0.10 | -0.08-0.28 |
| **MR-Egger** | -0.02 | -0.14-0.12 | 0.10 | -0.87-1.06 | 0.04 | -0.13-0.22 | 0.17 | -1.06-1.40 |

We cannot verify that alleles reported by CardioGRAMplusC4D or MEGASTROKE have been correctly orientated, therefore we removed palindromic SNPs if the allele frequency was close to 50%. Estimates are expressed as Log OR per-1-log unit increase in ever use of cannabis. Conventional MR was pooled effect across SNPs using fixed-effect with inverse variance weighted meta-analysis. No evidence against the null hypothesis of no directional pleiotropy of the genetic markers using MR-Egger intercept was found for overall and restricted SNPs (P-value for pleiotropy=0.753 and =0.684 for CAD; p-value=0.855 and =0.921 for IS, respectively). *for significant p-value (p-value<0.05)

**Supplementary Table 10.** Comparison of conventional MR, MR-Egger and weighted median MR causal effect estimates of cannabis use on ASCVD using cannabis use disorder as modifiable exposure

| | **Coronary artery disease** | | **Acute ischemic stroke** | |
|---|---|---|---|---|
| | **103 SNPs (p-value<5x10$^{-5}$)** | | **106 SNPs (p-value<5x10$^{-5}$)** | |
| **Analysis** | **Causal effect estimate** | **95% CI** | **Causal effect estimate** | **95% CI** |
| **Conventional MR** | 0.02 | -0.002-0.04 | 0.03 | -0.0004-0.06 |
| **MR-Egger** | 0.03 | -0.05-0.08 | 0.06 | -0.0006-0.12* |
| **Weighted median MR** | 0.007 | -0.03-0.04 | 0.02 | -0.01-0.06 |

We selected SNPs with p-value that were associated with cannabis use disorder (p-value<5x10$^{-5}$) and in low linkage-disequilibrium with other SNPs ($R^2$< 0.001) within a clumping distance of 10,000 kb. Palindromic SNPs with intermediate minor allele frequency were removed because we cannot assumed that all alleles were correctly reported in the positive strand in cannabis use disorder GWAS. A more restricted threshold with p-value <5x10$^{-8}$ was initially computed but only 2 SNPs remained, which was not sufficient for two-sample mendelian randomization analysis. Estimates are expressed as Log OR per-1-log unit increase in ever use of cannabis. Conventional MR was pooled effect across SNPs using fixed-effect with inverse variance weighted meta-analysis. *for significant p-value (p-value<0.05). There was evidence of directional pleiotropy for IS with a p-value=0.048 using MR-Egger intercept (p-value=0.116 for CAD).

**Supplementary Table 11.** Excluded studies from systematic reviews literature research

| Systematic reviews identified by the literature research | Reasons for exclusion |
|---|---|
| PMID 28432636 | 81 case reports study<br>12 case series study<br>2 experimental studies<br>3 clinical trial without outcome of interest<br>1 forum discussion<br>4 descriptive analyses of exposure population only<br>8 with inappropriate outcome<br>2 with inappropriate exposure (trigger instead of lifetime use or other cannabis as co-variable) |
| PMID 29357394 | 1 study was retracted<br>2 with inappropriate exposure (trigger instead of lisetime use)<br>5 with inappropriate outcome |
| PMID 34001774 | 2 case series study<br>4 lack of association analysis or descriptive analysis without controls<br>4 with inappropriate outcome<br>1 with inappropriate exposure (trigger instead of lifetime) |
| PMID 33636088 | 4 descriptive study<br>2 with inappropriate exposure (trigger instead of lifetime use<br>3 with inappropriate outcome (Moon cardiovascular event leading surgical procedure) |

**Supplementary Table 12**. Literature research' strategies

| **Steps of literature researchs** | **Exposure terms** | **Outcome terms** | **Others terms** |
|---|---|---|---|
| Research of meta-analysis | ("cannabis*" [MeSH] OR "marijuana*"[MeSH] OR "cannabinoids" OR "delta-9-tetrahydrocannabinol" OR "cannabidiol" OR "cannabinol" OR "sativex" OR "hash" OR "ganka" OR "weed" OR "hemp" OR "THC" OR "CBD") | ("Cardiovascular Diseases" [MeSH] OR "Myocardial Infarction"[Mesh] or "Stroke" [Mesh] OR "Cardiac Disease" OR "Heart Disease" OR "Vascular Disease" OR "Acute myocardial ischemia" OR "Myocardial*" OR "Acute Coronary Syndrome" OR "Angor" OR "Coronary heart disease" OR "Cardiac Ischemia" OR "Atherosclerosis" OR "Cerebrovascular disease" OR "Ischemic transient attack" OR "Cardiac Arrest" OR "Heart Arrest" OR "Cardiovascular death") | "humans"[MeSH]<br><br>AND<br><br>"Meta-Analysis" [Publication Type] |
| Reasearch of systematic reviews | Similar | Similar | "humans"[MeSH]<br>AND<br>"Systematic Review" [Publication Type] |
| Research for pospective or retrospectives observational studies* | Similar | Similar | "humans"[MeSH]<br>AND<br>("Cohort Studies"[Mesh] OR "Proportional Hazards Models"[Mesh] OR "prospective" OR "retrospective") |

*We used a Year timeline for this research from 2016 to august 2022, because the older systematic review included in our comprehensive literature analysis was in 2016.

### References

1. Ross S, Gerstein HC, Eikelboom J, Anand SS, Yusuf S, Paré G. Mendelian randomization analysis supports the causal role of dysglycaemia and diabetes in the risk of coronary artery disease. Eur Heart J. 2015 Jun 14;36(23):1454–62.

2. Vaucher HB Nadine Hausler, David Nanchen, Marie Méan, Pedro Marques Vidal, Julien. Comparison of Swiss and European risk algorithms for cardiovascular prevention in Switzerland - Hadrien Beuret, Nadine Hausler, David Nanchen, Marie Méan, Pedro Marques-Vidal, Julien Vaucher,. European Journal of Preventive Cardiology [Internet]. 2020 Feb 23 [cited 2020 Apr 7]; Available from: https://journals.sagepub.com/doi/full/10.1177/2047487320906305?rfr_dat=cr_pub%3Dpubmed&url_ver=Z39.88-2003&rfr_id=ori%3Arid%3Acrossref.org&journalCode=cprc

3. European Monitoring Centre for Drugs and Drug Addiction. European drug report 2021: trends and developments. [Internet]. LU: Publications Office; 2021 [cited 2022 Jul 4]. Available from: https://data.europa.eu/doi/10.2810/18539

4. Shim H, Chasman DI, Smith JD, Mora S, Ridker PM, Nickerson DA, Krauss RM, Stephens M. A multivariate genome-wide association analysis of 10 LDL subfractions, and their response to statin treatment, in 1868 Caucasians. PLoS One. 2015;10(4):e0120758.

5. Pierce BL, Burgess S. Efficient design for Mendelian randomization studies: subsample and 2-sample instrumental variable estimators. Am J Epidemiol. 2013 Oct 1;178(7):1177–84.

6. Firmann M, Mayor V, Vidal PM, Bochud M, Pécoud A, Hayoz D, Paccaud F, Preisig M, Song KS, Yuan X, Danoff TM, Stirnadel HA, Waterworth D, Mooser V, Waeber G, Vollenweider P. The CoLaus study: a population-based study to investigate the epidemiology and genetic determinants of cardiovascular risk factors and metabolic syndrome. BMC Cardiovasc Disord. 2008 Mar 17;8:6.

7. Johnson EC, Demontis D, Thorgeirsson TE, Walters RK, Polimanti R, Hatoum AS, Sanchez-Roige S, Paul SE, Wendt FR, Clarke TK, Lai D, Reginsson GW, Zhou H, He J, Baranger DAA, Gudbjartsson DF, Wedow R, Adkins DE, Adkins AE, Alexander J, Bacanu SA, Bigdeli TB, Boden J, Brown SA, Bucholz KK, Bybjerg-Grauholm J, Corley RP, Degenhardt L, Dick DM, Domingue BW, Fox L, Goate AM, Gordon SD, Hack LM, Hancock DB, Hartz SM, Hickie IB, Hougaard DM, Krauter K, Lind PA, McClintick JN, McQueen MB, Meyers JL, Montgomery GW, Mors O, Mortensen PB, Nordentoft M, Pearson JF, Peterson RE, Reynolds MD, Rice JP, Runarsdottir V, Saccone NL, Sherva R, Silberg JL, Tarter RE, Tyrfingsson T, Wall TL, Webb BT, Werge T, Wetherill L, Wright MJ, Zellers S, Adams MJ, Bierut LJ, Boardman JD, Copeland WE, Farrer LA, Foroud TM, Gillespie NA, Grucza RA, Harris KM, Heath AC, Hesselbrock V, Hewitt JK, Hopfer CJ, Horwood J, Iacono WG, Johnson EO, Kendler KS, Kennedy MA, Kranzler HR, Madden PAF, Maes HH, Maher BS, Martin NG, McGue M, McIntosh AM, Medland SE, Nelson EC, Porjesz B, Riley BP, Stallings MC, Vanyukov MM, Vrieze S, Davis LK, Bogdan R, Gelernter J, Edenberg HJ, Stefansson K, Børglum AD, Agrawal A. A large-scale genome-wide association study meta-analysis of cannabis use disorder. Lancet Psychiatry. 2020 Dec;7(12):1032–45.

8. Pasman JA, Verweij KJH, Gerring Z, Stringer S, Sanchez-Roige S, Treur JL, Abdellaoui A, Nivard MG, Baselmans BML, Ong JS, Ip HF, van der Zee MD, Bartels M, Day FR, Fontanillas P, Elson SL, 23andMe Research Team, de Wit H, Davis LK, MacKillop J, Substance Use Disorders Working Group of the Psychiatric Genomics Consortium, International Cannabis Consortium, Derringer JL, Branje SJT, Hartman CA, Heath AC, van Lier PAC, Madden PAF, Mägi R, Meeus W, Montgomery GW, Oldehinkel AJ, Pausova Z, Ramos-Quiroga JA, Paus T,

Ribases M, Kaprio J, Boks MPM, Bell JT, Spector TD, Gelernter J, Boomsma DI, Martin NG, MacGregor S, Perry JRB, Palmer AA, Posthuma D, Munafò MR, Gillespie NA, Derks EM, Vink JM. GWAS of lifetime cannabis use reveals new risk loci, genetic overlap with psychiatric traits, and a causal influence of schizophrenia. Nat Neurosci. 2018 Sep;21(9):1161–70.

9. Reis JP, Auer R, Bancks MP, Goff DC, Lewis CE, Pletcher MJ, Rana JS, Shikany JM, Sidney S. Cumulative Lifetime Marijuana Use and Incident Cardiovascular Disease in Middle Age: The Coronary Artery Risk Development in Young Adults (CARDIA) Study. Am J Public Health. 2017 Apr;107(4):601–6.

10. Chami T, Kim CH. Cannabis Abuse and Elevated Risk of Myocardial Infarction in the Young: A Population-Based Study. Mayo Clin Proc. 2019 Aug;94(8):1647–9.

11. Phillips KT, Pedula KL, Choi NG, Tawara KAK, Simiola V, Satre DD, Owen-Smith A, Lynch FF, Dickerson J. Chronic health conditions, acute health events, and healthcare utilization among adults over age 50 in Hawai'i who use cannabis: A matched cohort study. Drug Alcohol Depend. 2022 May 1;234:109387.

12. Patel RS, Manocha P, Patel J, Patel R, Tankersley WE. Cannabis Use Is an Independent Predictor for Acute Myocardial Infarction Related Hospitalization in Younger Population. J Adolesc Health. 2020 Jan;66(1):79–85.

13. Burt JR, Agha AM, Yacoub B, Zahergivar A, Pepe J. Marijuana use and coronary artery disease in young adults. PLoS One. 2020;15(1):e0228326.

14. Falkstedt D, Wolff V, Allebeck P, Hemmingsson T, Danielsson AK. Cannabis, Tobacco, Alcohol Use, and the Risk of Early Stroke: A Population-Based Cohort Study of 45 000 Swedish Men. Stroke. 2017 Feb;48(2):265–70.

15. Rumalla K, Reddy A, Mittal M. Recreational marijuana Use and acute ischemic stroke: A population-based analysis of hospitalized patients in the United States. Journal of the Neurological Sciences. 2016 Feb 1;364.

16. Westover AN, McBride S, Haley RW. Stroke in young adults who abuse amphetamines or cocaine: a population-based study of hospitalized patients. Arch Gen Psychiatry. 2007 Apr;64(4):495–502.

17. Vin-Raviv N, Akinyemiju T, Meng Q, Sakhuja S, Hayward R. Marijuana use and inpatient outcomes among hospitalized patients: analysis of the nationwide inpatient sample database. Cancer Med. 2017 Jan;6(1):320–9.

18. Sun Y, Liu B, Wallace RB, Bao W. Association of Cannabis Use With All-Cause and Cause-Specific Mortality Among Younger- and Middle-Aged U.S. Adults. Am J Prev Med. 2020 Dec;59(6):873–9.

19. Auger N, Paradis G, Low N, Ayoub A, He S, Potter BJ. Cannabis use disorder and the future risk of cardiovascular disease in parous women: a longitudinal cohort study. BMC Med. 2020 Nov 19;18(1):328.

20. Bowden J, Davey Smith G, Burgess S. Mendelian randomization with invalid instruments: effect estimation and bias detection through Egger regression. Int J Epidemiol. 2015 Apr;44(2):512–25.

21. Nikpay M, Goel A, Won HH, Hall LM, Willenborg C, Kanoni S, Saleheen D, Kyriakou T, Nelson CP, Hopewell JC, Webb TR, Zeng L, Dehghan A, Alver M, Armasu SM, Auro K, Bjonnes A, Chasman DI, Chen S, Ford I, Franceschini N, Gieger C, Grace C, Gustafsson S, Huang J, Hwang SJ, Kim YK, Kleber ME, Lau KW, Lu X, Lu Y, Lyytikäinen LP, Mihailov E,

Morrison AC, Pervjakova N, Qu L, Rose LM, Salfati E, Saxena R, Scholz M, Smith AV, Tikkanen E, Uitterlinden A, Yang X, Zhang W, Zhao W, de Andrade M, de Vries PS, van Zuydam NR, Anand SS, Bertram L, Beutner F, Dedoussis G, Frossard P, Gauguier D, Goodall AH, Gottesman O, Haber M, Han BG, Huang J, Jalilzadeh S, Kessler T, König IR, Lannfelt L, Lieb W, Lind L, Lindgren CM, Lokki ML, Magnusson PK, Mallick NH, Mehra N, Meitinger T, Memon F ur R, Morris AP, Nieminen MS, Pedersen NL, Peters A, Rallidis LS, Rasheed A, Samuel M, Shah SH, Sinisalo J, Stirrups KE, Trompet S, Wang L, Zaman KS, Ardissino D, Boerwinkle E, Borecki IB, Bottinger EP, Buring JE, Chambers JC, Collins R, Cupples LA, Danesh J, Demuth I, Elosua R, Epstein SE, Esko T, Feitosa MF, Franco OH, Franzosi MG, Granger CB, Gu D, Gudnason V, Hall AS, Hamsten A, Harris TB, Hazen SL, Hengstenberg C, Hofman A, Ingelsson E, Iribarren C, Jukema JW, Karhunen PJ, Kim BJ, Kooner JS, Kullo IJ, Lehtimäki T, Loos RJF, Melander O, Metspalu A, März W, Palmer CN, Perola M, Quertermous T, Rader DJ, Ridker PM, Ripatti S, Roberts R, Salomaa V, Sanghera DK, Schwartz SM, Seedorf U, Stewart AF, Stott DJ, Thiery J, Zalloua PA, O'Donnell CJ, Reilly MP, Assimes TL, Thompson JR, Erdmann J, Clarke R, Watkins H, Kathiresan S, McPherson R, Deloukas P, Schunkert H, Samani NJ, Farrall M, the CARDIoGRAMplusC4D Consortium. A comprehensive 1000 Genomes–based genome-wide association meta-analysis of coronary artery disease. Nat Genet. 2015 Oct;47(10):1121–30.

### Supplementary Figures

**Supplementary Figure 1.** Venn diagram showing the number of individuals and the overlap between the cannabis-GWAS and the CAD-GWAS

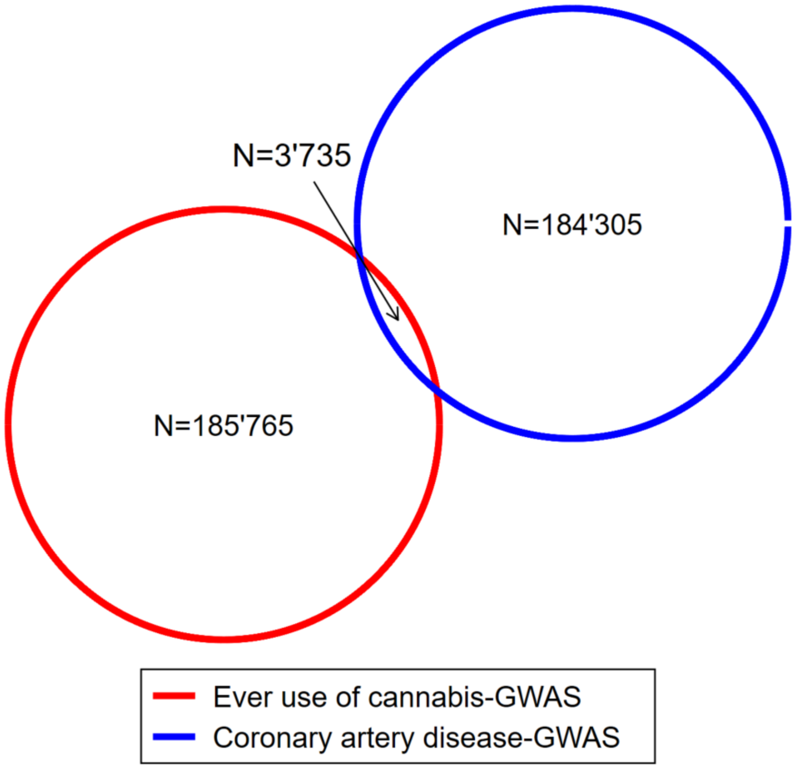

Based on the data provided in Pasman[8] et al and Nikpay et al[21], only the ECGUT study (Estonian Genome Center University of Tartu (N=3,735)) contributed to both GWAS.

**Supplementary Figure 2**. Flow chart of selection of studies included in our meta-analysis

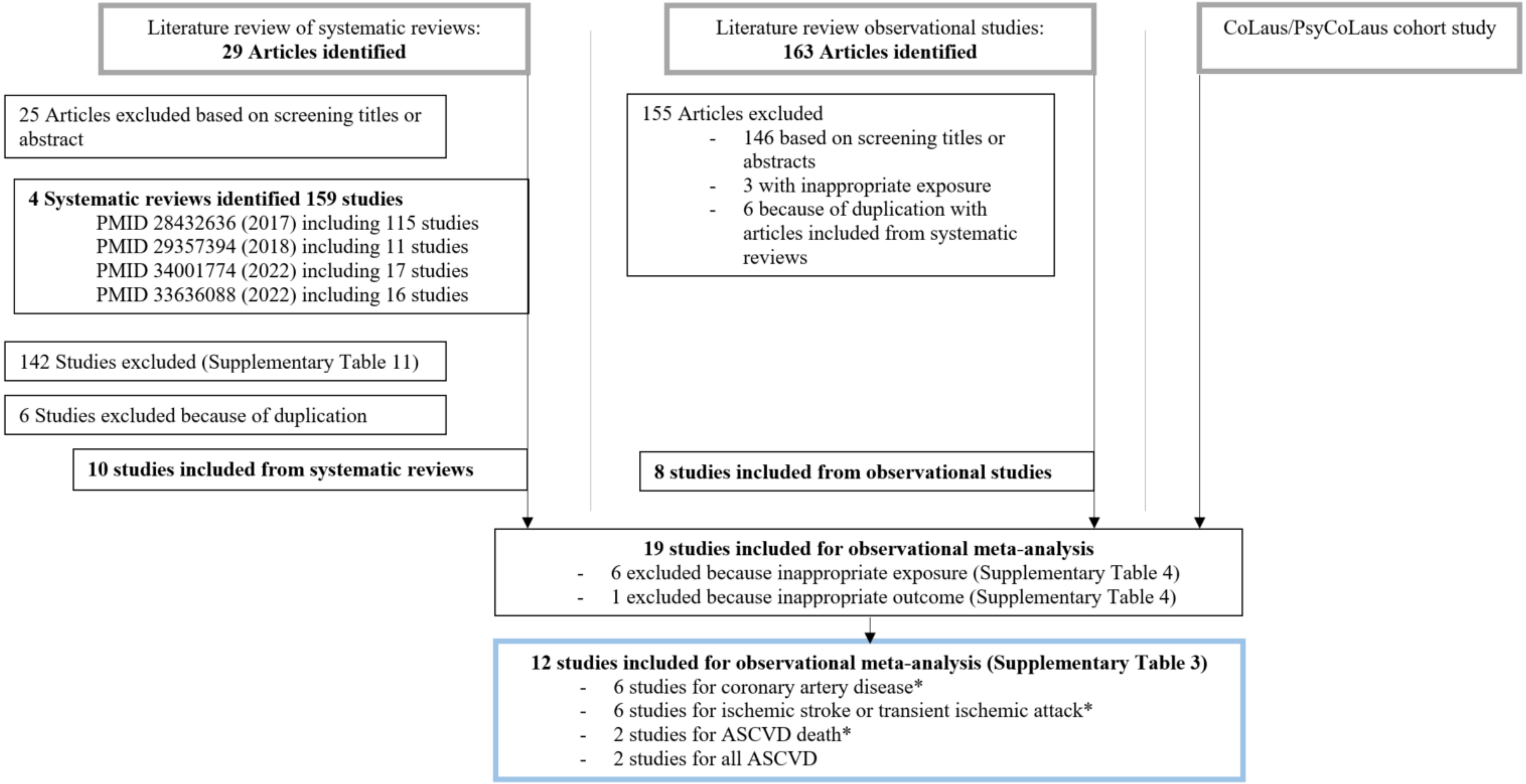

Details for research strategy can be found in Supplementary Table 12. *sum of subtotal studies are not equal to total studies because some studies accounts for different outcomes.

**Supplementary Figure 3**. Meta-analysis and forest plot of observational studies reporting an association between lifetime cannabis use and risk of coronary artery disease

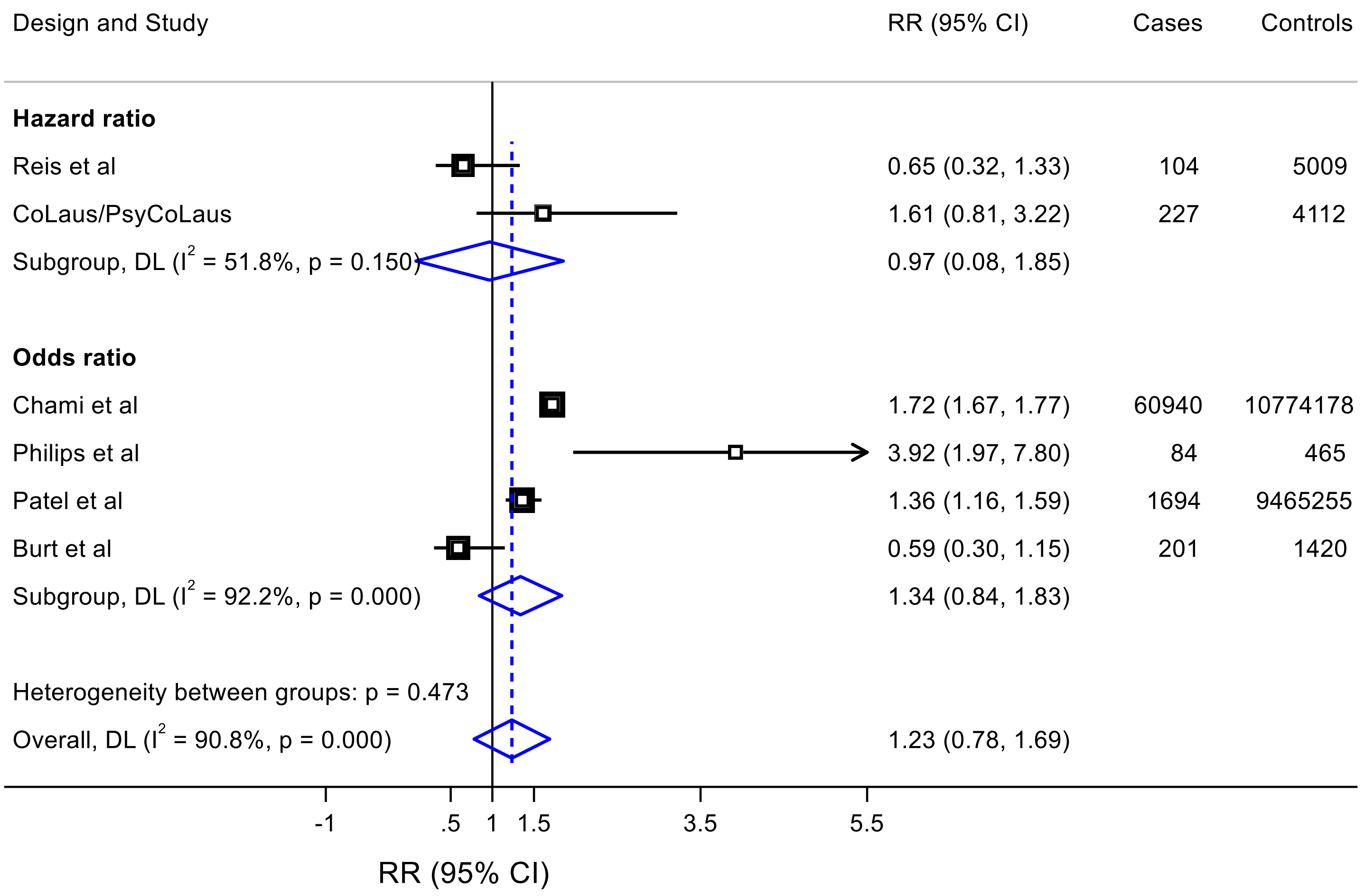

Random-effect meta-analysis was calculated using DerSimonian and Laird methods (DL). Boxes are scaled according to the weight of the study in the random-effect model.

**Supplementary Figure 4**. Meta-analysis and forest plot of observational studies reporting an association between lifetime cannabis use and risk of ischemic stroke

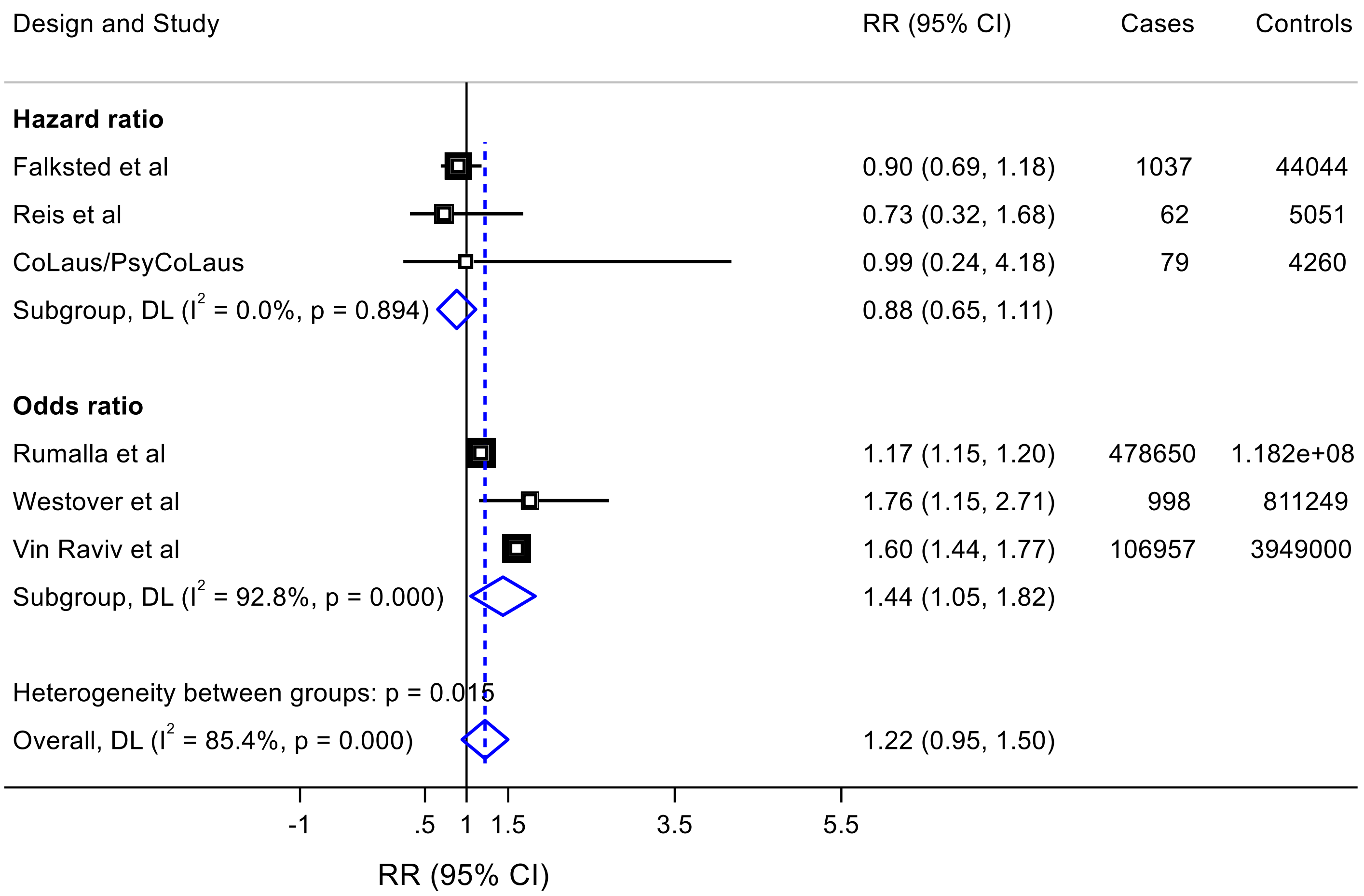

Random-effect meta-analysis was calculated using DerSimonian and Laird methods (DL). Boxes are scaled according to the weight of the study in the random-effect model.

**Supplementary Figure 5**. Meta-analysis and forest plot of prospective observational studies reporting an association between lifetime cannabis use and risk of arteriosclerotic cardiovascular disease

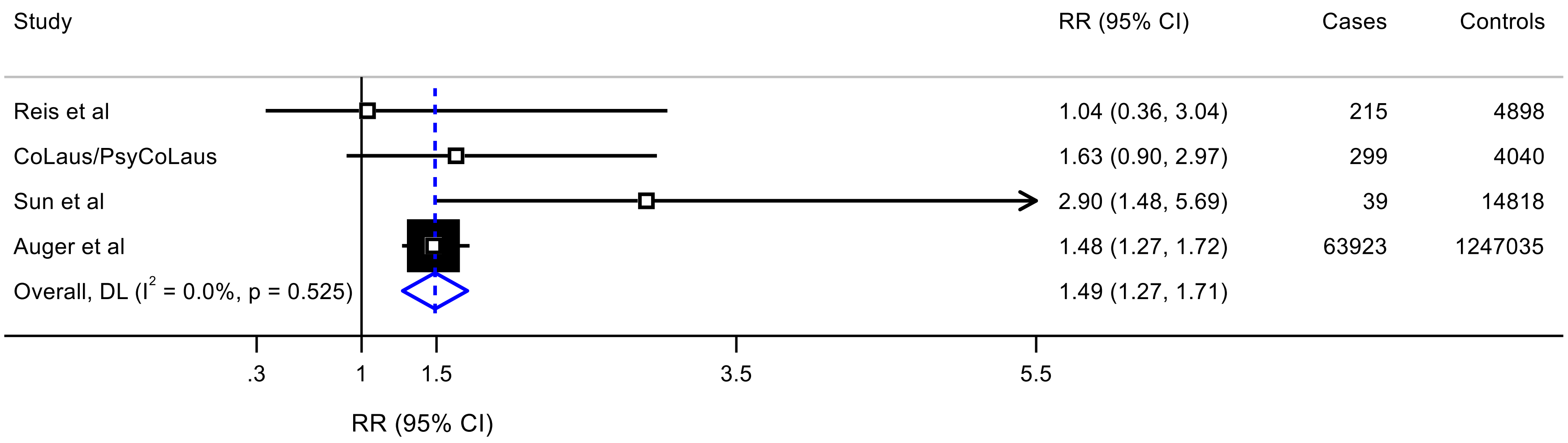

Random-effect meta-analysis was calculated using DerSimonian and Laird methods (DL). Boxes are scaled according to the weight of the study in the random-effect model.

**Supplementary Figure 6**. Pair-wise association plot of the SNPs associated with cannabis use and risk of CAD

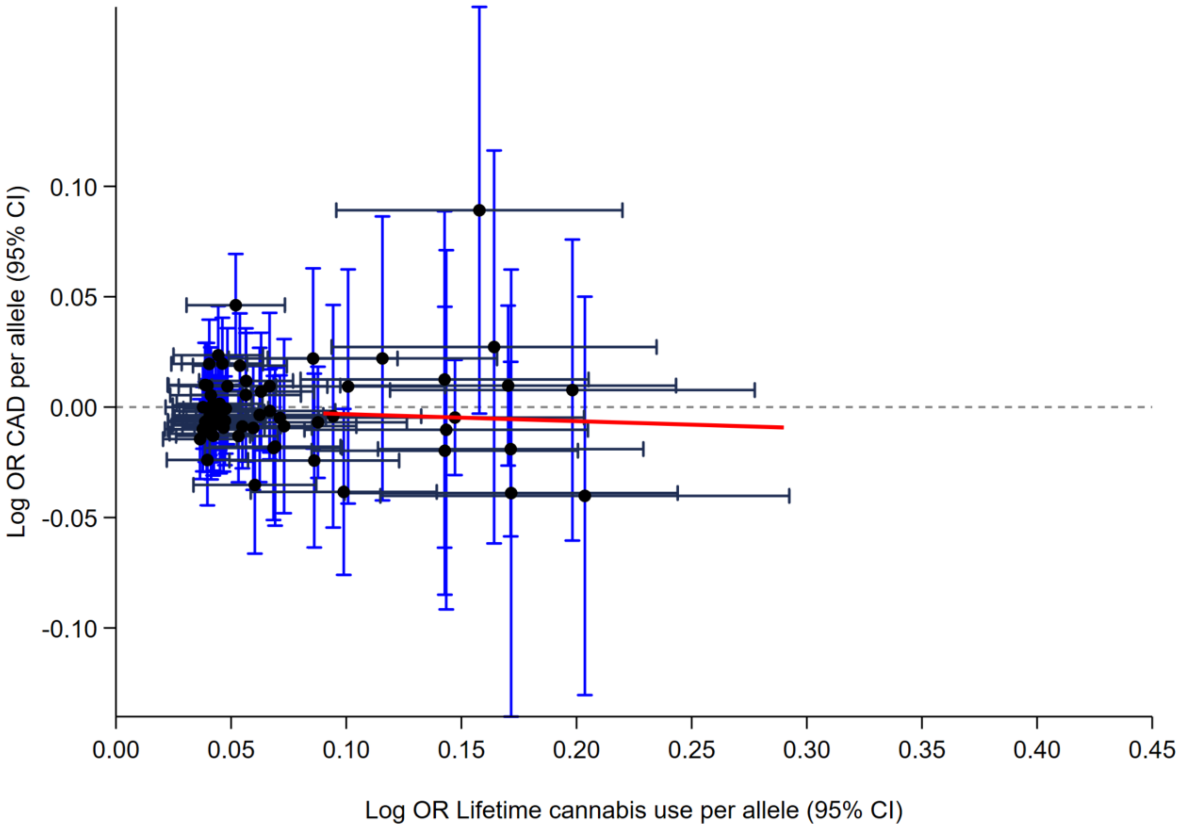

The red line represents the regression slope of the causal effects estimates (derived by the inverse-variance weighted approach as proposed by Bowden et al.).14

**Supplementary Figure 7**. Pair-wise association plot of the 10 SNPs associated with cannabis use and risk of IS

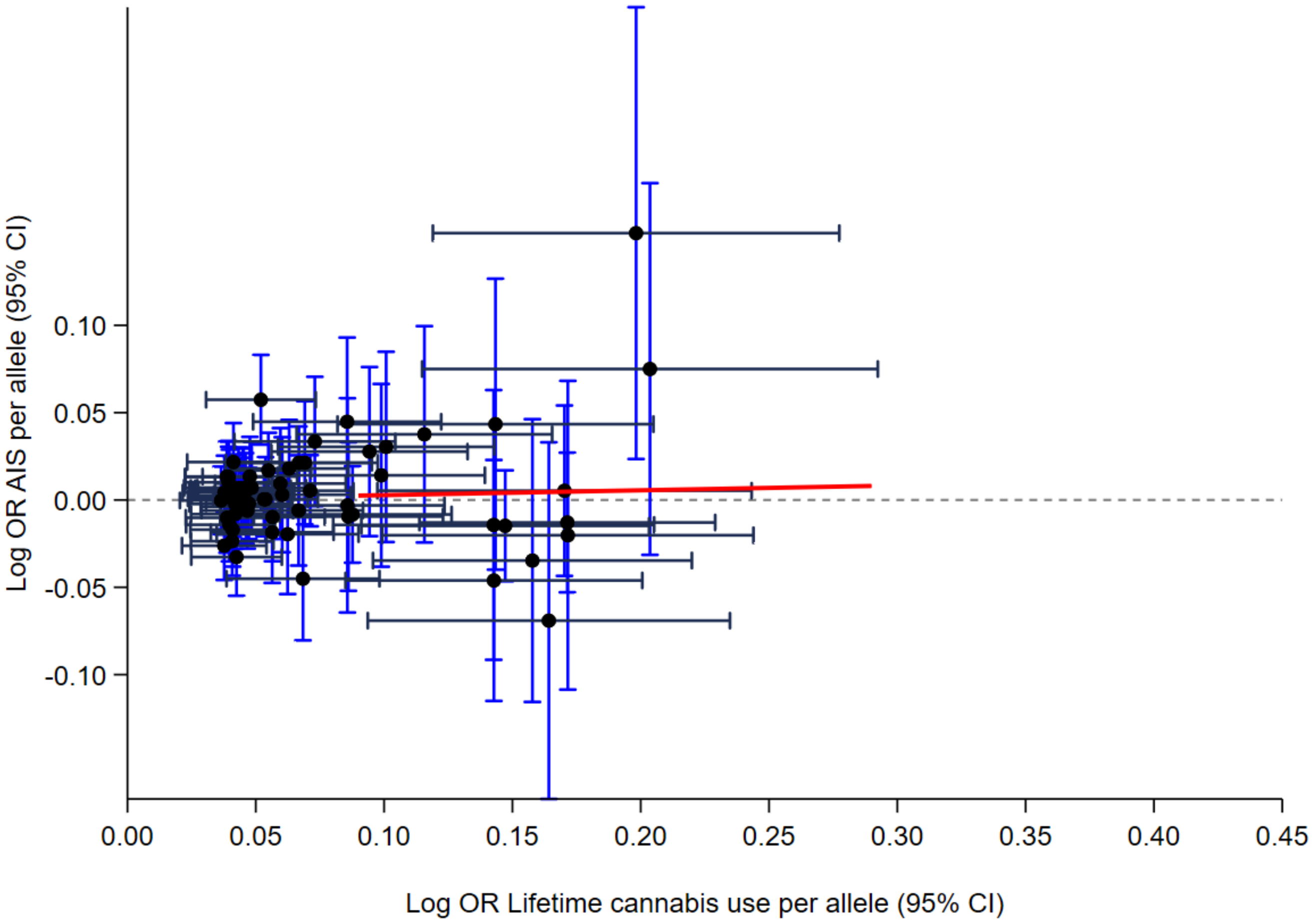

The red line represents the regression slope of the causal effects estimates (derived by the inverse-variance weighted approach as proposed by Bowden et al.).14

**Supplementary Figure 8.** Scatter plot of the genetic association with cannabis use against genetic association with coronary artery disease

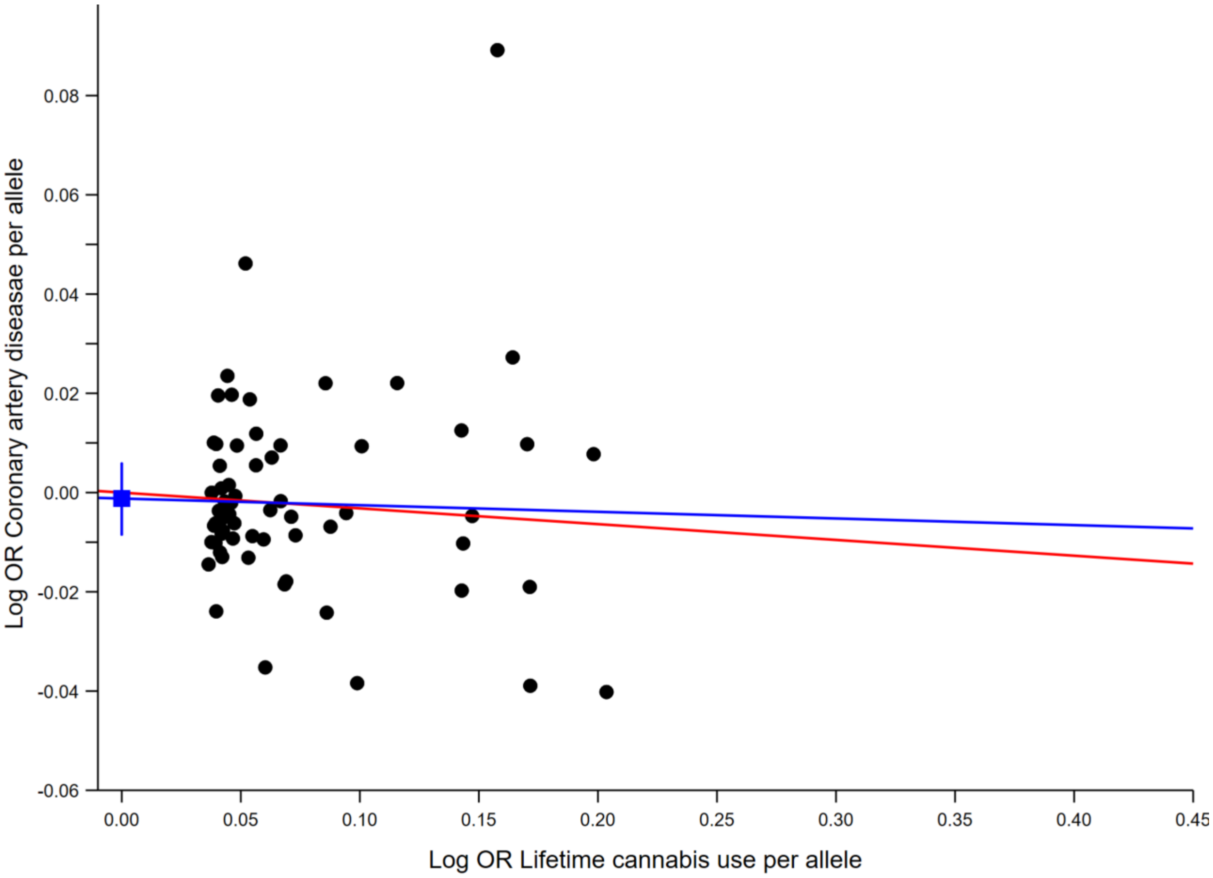

The conventional Mendelian randomization (Conventional MR in red), Egger Mendelian randomization (MR-Egger in blue) causal effects estimates are presented as regression slopes. The constant and its 95% CI (obtained by bootstrap resampling 10,000 times) derived from Egger regression are shown as the blue square and vertical bar, respectively.

**Supplementary Figure 9.** Scatter plot of the genetic association with cannabis use against genetic association with acute ischemic stroke

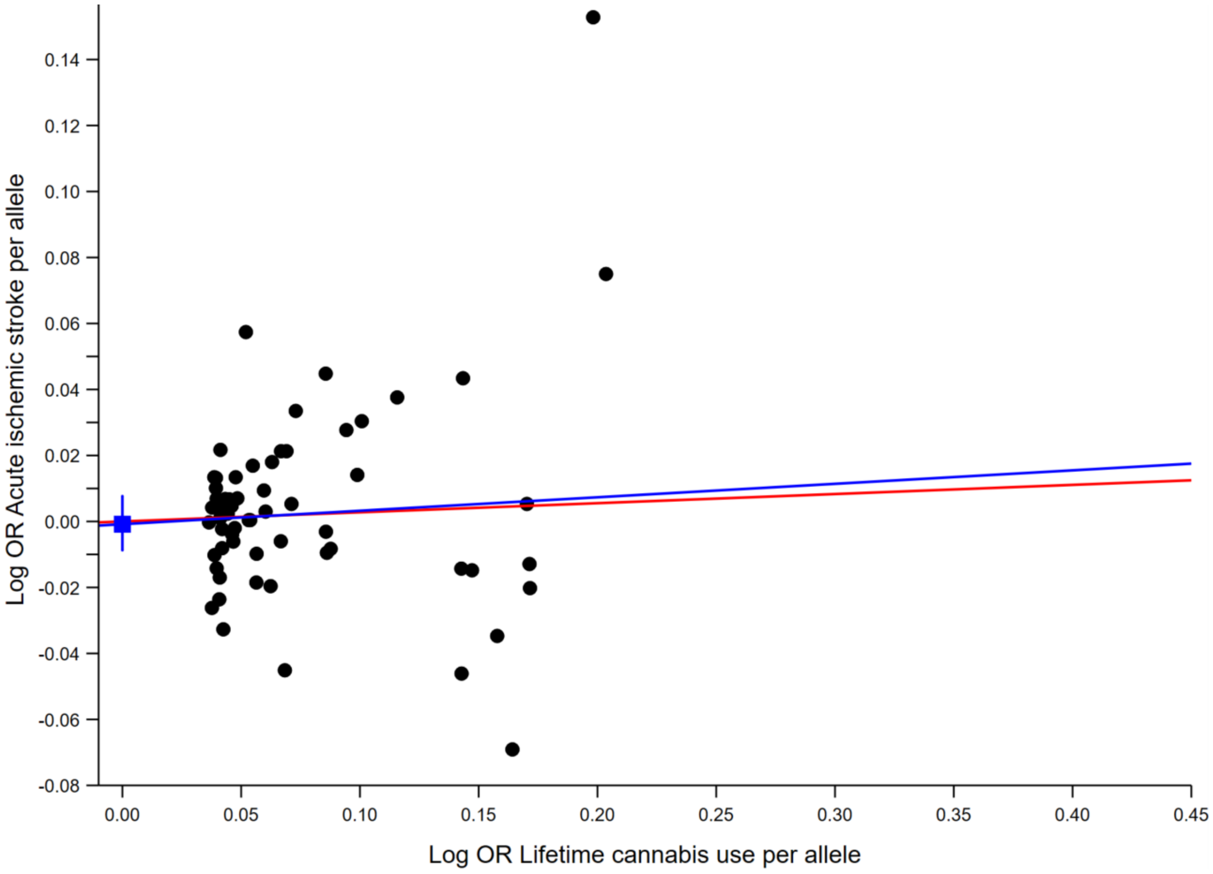

The conventional Mendelian randomization (Conventional MR in red), Egger Mendelian randomization (MR-Egger in blue) causal effects estimates are presented as regression slopes. The constant and its 95% CI (obtained by bootstrap resampling 10,000 times) derived from Egger regression are shown as the blue square and vertical bar, respectively.
